# Efficacy and safety of hydroxychloroquine as pre-and post-exposure prophylaxis and treatment of COVID-19. Systematic review and meta-analysis of blinded, placebo-controlled, randomized clinical trials

**DOI:** 10.1101/2021.06.12.21258831

**Authors:** Paulo Ricardo Martins-Filho, Lis Campos Ferreira, Luana Heimfarth, Adriano Antunes de Souza Araújo, Lucindo José Quintans-Júnior

## Abstract

**BACKGROUND:** Hydroxychloroquine (HCQ) is an anti-malarial and immunomodulatory drug considered a potential candidate for drug repurposing in COVID-19 due to their *in vitro* antiviral activity against SARS-CoV-2. Despite the potential antiviral effects and anti-inflammatory profile, the results based on clinical studies are contradictory and the quality of the decision-making process from meta-analyses summarizing the available evidence selecting studies with different designs and unblinded trials is limited. The aim of this study was to synthesize the best evidence on the efficacy and safety of HCQ as pre-and post-exposure prophylaxis and treatment of non-hospitalized and hospitalized patients with COVID-19.

**METHODS:** Searches for studies were performed in PubMed, Web of Science, Embase, Lilacs, the website ClinicalTrials.gov and the preprint server medRxiv from January 1, 2020 to May 17, 2021. The following elements were used to define eligibility criteria: (1) Population, individuals at high-risk of exposure to SARS-CoV-2 (pre-exposure), individuals who had close contact with a positive or probable case of COVID-19 (post-exposure), non-hospitalized patients with COVID-19 and hospitalized patients with COVID-19; (2) Intervention, HCQ; (3) Comparison, placebo; (4) Outcomes: incidence of SARS-CoV-2 infection, need for hospitalization, length of hospital stay, need for invasive mechanical ventilation (MV), death, and adverse events; and (5) Study type, blinded, placebo-controlled, randomized clinical trials (RCTs). Risk of bias was judged according to the Cochrane guidelines for RCTs. Treatment effects were reported as relative risk (RR) for dichotomous variables and mean difference (MD) for continuous variables with 95% confidence intervals (CI). We used either a fixed or random-effects model to pool the results of individual studies depending on the presence of heterogeneity. The GRADE system was used to evaluate the strength of evidence between use of HCQ and the outcomes of interest.

**RESULTS:** Fourteen blinded, placebo-controlled RCTs were included in the meta-analysis. Four trials used HCQ as a prophylactic medication pre-exposure to COVID-19, two as a prophylactic medication post-exposure to COVID-19, three as treatment for non-hospitalized patients, and five as treatment for hospitalized patients with COVID-19. We found no decreased risk of SARS-CoV-2 infection among individuals receiving HCQ as pre-exposure (RR = 0.90; 95% CI 0.46 to 1.77) or post-exposure (RR = 0.96; 95% CI 0.72 to 1.29) prophylaxis to prevent COVID-19. There is no decreased risk of hospitalization for outpatients with SARS-CoV-2 infection (RR = 0.64; 95% CI 0.33 to 1.23) and no decreased risk of MV (RR = 0.81; 95% CI 0.49 to 1.34) and death (RR = 1.05; 95% CI 0.62 to 1.78) among hospitalized patients with COVID-19 receiving HCQ. The certainty of the results on the lack of clinical benefit for HCQ was rated as moderate. Moreover, our results demonstrated an increased risk for any adverse events and gastrointestinal symptoms among those using HCQ.

**CONCLUSION:** Available evidence based on the results of blinded, placebo-controlled RCTs showed no clinical benefits of HCQ as pre-and post-exposure prophylaxis and treatment of non-hospitalized and hospitalized patients with COVID-19.

## 1 INTRODUCTION

Chloroquine (CQ) and hydroxychloroquine (HCQ) are anti-malarial and immunomodulatory drugs that have been suggested for the prevention and treatment of coronavirus disease 2019 (COVID-19) due to their *in vitro* antiviral activity against the severe acute respiratory syndrome coronavirus-2 (SARS-CoV-2)^1,2^ and the potential for suppressing the release of proinflammatory cytokines.^3^ On March 20, 2020, promising results of HCQ in clearing viral nasopharyngeal carriage of SARS-CoV-2 in hospitalized patients with COVID-19 were described in a small open-label non-randomized clinical trial (RCT).^4^ These drugs are characterized as diprotic weak bases and can elevate endolysosomal pH inhibiting the virus/cell membrane fusion.^5^ Despite the potential antiviral effects and anti-inflammatory profile, the results based on clinical studies for patients with COVID-19 are contradictory and an increased risk for severe adverse events has been found for those treated with HCQ.^6^

Recent meta-analyses pooling results of open-label and blinded clinical trials have shown no clinical benefit of anti-malarial drugs on prophylaxis^7^ and treatment of COVID-19.^8^ Contrasting findings were described in a meta-analysis of clinical reports that showed improvement in clinical and virological outcomes for patients using CQ.^9^ Moreover, an evidence synthesis based on observational studies found a 7% to 33% reduced mortality in hospitalized patients with COVID-19 using lower doses of HCQ.^10^ Facing the inconsistent evidence on the effects of HCQ to prevent and treat COVID-19 and the increased risk of adverse events, the World Health Organization (WHO) has not recommended the use of HCQ in COVID-19 ^11^. Although the lack of rigorous evidence for efficacy, the politicization of the COVID-19 treatment in some countries and scientific denial have been important factors in promoting interest in use of this drug.^12^

The best evidence synthesis to assess treatment effects can be obtained through the identification, critical appraisal, and summary of results from blinded, placebo-controlled RCTs considered the gold standard in clinical research. Summarizing the available evidence selecting studies with different designs and unblinded trials may compromise the validity of the meta-analysis and the quality of the decision-making process. The aim of this study was to synthesize the best evidence on the efficacy and safety of HCQ as pre-and post-exposure prophylaxis and treatment of non-hospitalized and hospitalized patients with COVID-19.

## 2 METHODS

### 2.1 Search strategy

Searches for studies were performed in PubMed, Web of Science, Embase, Lilacs, the website ClinicalTrials.gov and the preprint server medRxiv from January 1, 2020 to May 17, 2021. The search was limited to studies published in full-text versions, without language restriction. In the ClinicalTrials.gov, only completed studies with results were analyzed. The reference lists of all eligible studies and reviews were scanned to identify additional studies for inclusion. The structured search strategies used for each database with specific filters and grey-literature were detailed in the supplementary file.

### 2.2 Study selection and eligibility criteria

Two reviewers (P.R.M.-F. and L.C.F.) independently screened the search results and identified studies that were potentially relevant based on their title and abstract. Relevant studies were read in full and selected according to eligibility criteria. Disagreements between the two reviewers were resolved by consensus.

The following elements were used to define eligibility criteria:

1. Population: Individuals at high-risk of exposure to SARS-CoV-2 (pre-exposure), individuals who had close contact with a positive or probable case of COVID-19 (post-exposure), non-hospitalized patients with COVID-19 and hospitalized patients with COVID-19.
2. Intervention: HCQ. Trials that tested drug associations were excluded.
3. Comparison: Placebo.
4. Outcomes:
  ▪ Pre- and post-exposure prophylaxis: incidence of SARS-CoV-2 infection and any adverse events.
  ▪ Non-hospitalized patients with COVID-19: need for hospitalization, death, and any adverse events.
  ▪ Hospitalized patients with COVID-19: length of hospital stay, need for invasive mechanical ventilation (MV), death, and any adverse events.
5. Study type: blinded, placebo-controlled, RCTs. Eligible studies must report at least 1 of the outcomes of interest. Potential overlapping populations, open-label trials, and observational studies were excluded.

### 2.3 Data extraction

Two authors (P.R.M.-F. and L.C.F.) extracted the data from included studies and crosschecked them for accuracy. Using a standardized data extraction sheet, the following information were extracted from the studies: registry of study protocol, demographic characteristics of study participants, pre-existing medical conditions, treatment arms, HCQ protocol, concomitant medications, follow-up duration, and outcome data. We also extracted data on serious adverse events, QTc interval >500ms and other cardiac manifestations, gastrointestinal symptoms (vomiting, diarrhea, and abdominal pain), skin reaction (rash), headache, and neurologic reactions (irritability, dizziness, vertigo, and seizures).

### 2.4 Risk of bias assessment

Risk of bias was judged according to the Cochrane guidelines for RCTs.^13^ The following domains were evaluated: sequence generation and allocation concealment (selection bias), blinding of participants and personnel (performance bias), blinding of outcome assessment (detection bias), incomplete outcome data (attrition bias), selective outcome reporting (reporting bias), sample size calculation, power analysis, and early stopping for futility (operational bias), outcome measurements (information bias), and the authors’ financial or non-financial conflicts of interest that could appear to affect the judgment of research team when designing, conducting, or reporting study. Self-reported diagnosis of COVID-19 was classified as a high risk of information bias. Studies using real-time reverse transcription polymerase chain reaction (RT-PCR) to detect SARS-CoV-2 or, if testing was limited, provided a clinical diagnosis based on COVID-19-related symptoms and epidemiological data were considered as having a low risk of bias.

### 2.5 Data synthesis

Treatment effects were reported as relative risk (RR) for dichotomous variables (incidence of SARS-CoV-2 infection, need for hospitalization, need for MV, death, and adverse events) and mean difference (MD) for continuous variables (length of hospital stay) with 95% confidence intervals (CI). To calculate MD, means and standard deviations (SD) were obtained for each study group. If the means and SD were not directly reported in the publication, indirect methods of extracting estimates were used.^14^ A negative effect size indicated that HCQ was beneficial in reducing the length of stay for hospitalized patients with COVID-19. To calculate the RR, the number of events and individuals in each treatment group were extracted. For studies using three or more different dosages of HCQ, we combined all treatment groups to a single large group.

We used either a fixed or random-effects model to pool the results of individual studies depending on the presence of heterogeneity. Statistical heterogeneity was quantified by the I^2^ index using the following interpretation: 0%, no between-study heterogeneity; <50%, low heterogeneity; 50–75%, moderate heterogeneity; > 75%, high heterogeneity. ^15^ In the case of heterogeneity, we used the random-effects model, otherwise, the fixed-effects model was used. The results of the meta-analysis for each outcome were presented according to the population characteristics and therapeutic proposal (pre-exposure prophylaxis, post-exposure prophylaxis, treatment of non-hospitalized individuals and treatment of hospitalized individuals). We also provided an additional analysis on the overall risk for any and serious adverse events, QTc interval >500ms and other cardiac manifestations, gastrointestinal symptoms, skin rash, headache, and neurologic reactions for individuals receiving HCQ.

Although funnel plots may be useful tools in investigating small study effects in meta-analyses, they have limited power to detect such effects when there are few studies.^16^ Therefore, because we had only a small number of included studies according to the population characteristics and use of HCQ, we did not perform a funnel plot analysis. Forest plots were used to present the effect sizes and the 95% CI, and a 2-tailed p < 0.05 was used to determine significance. Analyses were conducted using Review Manager, version 5.3 (Cochrane IMS).

### 2.6 Grading the strength of evidence

We graded the strength of evidence for the association between use of HCQ and the outcomes of interest as high, moderate, low, or very-low using the Grading of Recommendations Assessment, Development, and Evaluation (GRADE) rating system.^17,18^ In the GRADE system, RCTs begin as high-quality evidence but may be downrated according to the risk of bias assessment, inconsistency, indirectness, imprecision in the results, and publication bias.^19^ Certainty is uprated for estimates with large (RR > 2.0 or RR < 0.5) or very-large (RR > 5.0 or RR < 0.2) magnitude of effect.

Quality of evidence was lowered when the proportion of information from studies at high risk of bias was sufficient to affect the interpretation of results. Evidence of inconsistency included important variations in point estimates, no overlapping of confidence intervals, inconsistency in direction of effect, and considerable between-study heterogeneity (I^2^ > 75%). Reasons of indirectness included differences in study populations and interventions, use of surrogate outcomes, and inadequate follow-up time. The width of CI for the pooled estimates and the optimal information size (OIS) were analyzed for imprecision.^20^ OIS was calculated using the following formula^21^: OIS = (4.(Z _1-α_ + Z_1-β_)^2^.P.(1-P)/d^2^).(1/1-I^2^)), where α = 5%, β = 80%, P = control group risk, δ = difference between control group risk and intervention group risk, and I^2^ = between-study heterogeneity.

Although the funnel plot asymmetry was not evaluated, we reduced the potential for publication bias planning a comprehensive search including grey-literature without restrictions. In this criterion, we analyzed discrepancies in findings between peer-reviewed and non-peer reviewed publications and the influence of small trials (< 100 patients per arm) on estimated treatment effects. The influence of non-peer reviewed publications and small trials on the pooled estimates was analyzed using a “leave-one-out” sensitivity approach.^22^

## 3 RESULTS

Search strategy yielded 2871 potentially relevant records. After screening of titles and abstracts and evaluation of completed trials retrieved from ClinicalTrials.gov, 33 full-text articles were assessed for eligibility and 14 blinded, placebo-controlled RCTs^23–36^ were included in the meta-analysis. No trials evaluating the efficacy and safety of CQ in COVID-19 were found. Of the included trials, four^33–36^ used HCQ as a prophylactic medication pre-exposure to COVID-19, two^31,32^ as a prophylactic medication post-exposure to COVID-19, three^28–30^ as treatment for non-hospitalized patients, and five^23–27^ as treatment for hospitalized patients with COVID-19. A flow diagram of the study selection process and specific reasons for exclusion are detailed in Figure 1.

**Figure 1.**
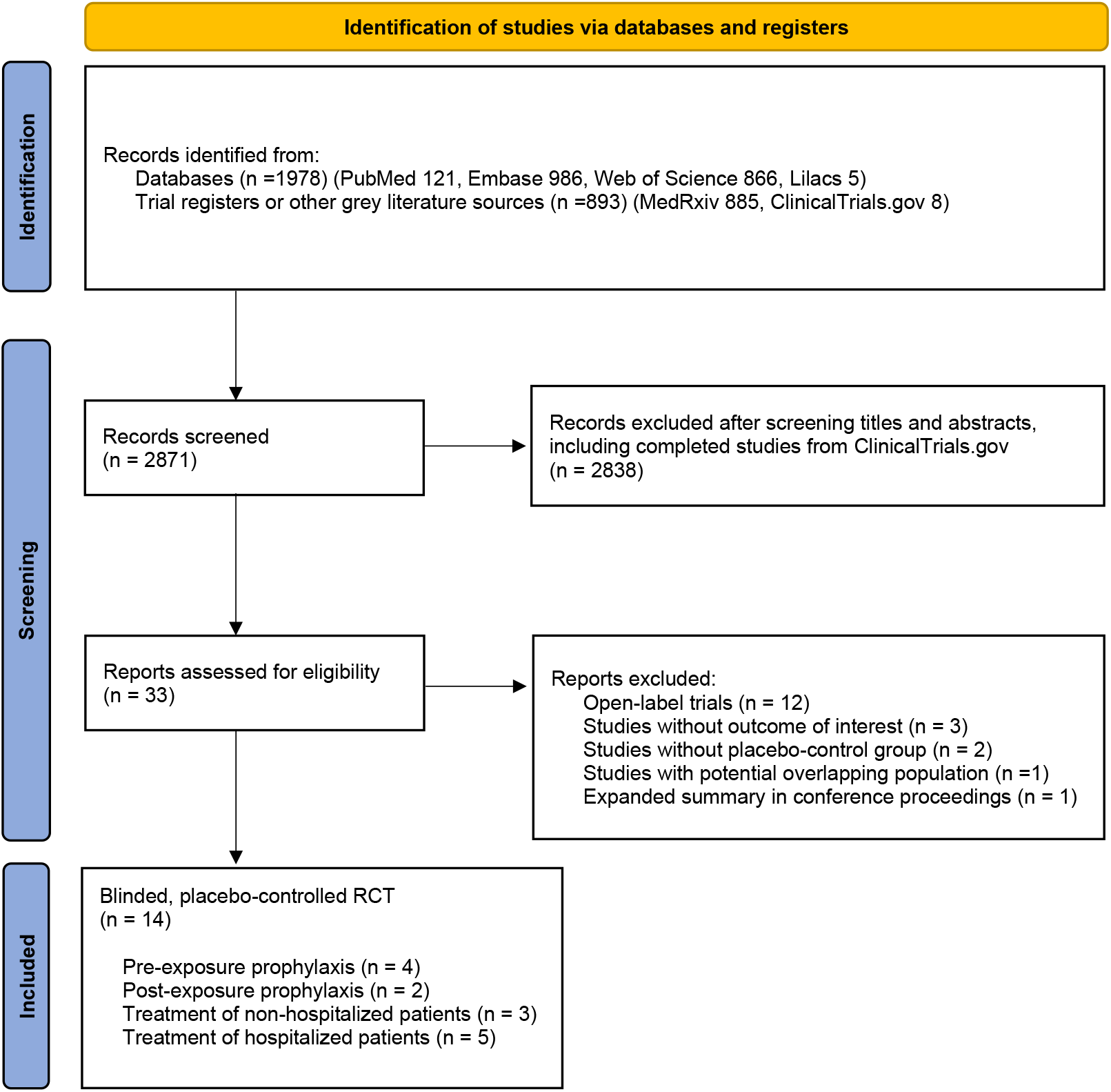
PRISMA flow chart of studies screened and included.

### 3.1 HCQ as pre-exposure prophylaxis to prevent SARS-CoV-2 infection

Population included healthcare workers in the USA, Canada, Mexico, and Pakistan at high-risk of exposure to SARS-CoV-2. Trials enrolled individuals with no history of SARS-CoV-2 infection and no symptoms suggestive of COVID-19. All trial protocols were registered on ClinicalTrials.gov, had a parallel design, and were classified as Phase 2 or Phase 3. Dosage regimens of HCQ were different between studies and outcomes were assessed up to 8 or 12 weeks (Table S1; supplementary file). Most studies had a low risk for selection, performance, detection, attrition, and information bias. One study was designed as a single blinded RCT and classified as having a high risk for detection bias.^34^ There was potential risk for reporting bias of some outcomes and a high risk for operational bias in all studies (Figure S1; supplementary file).

The four blinded, placebo-controlled RCTs enrolled a total of 1942 healthcare workers, 1271 in the HCQ group and 671 in the placebo group. Individual results showed no clinical benefit on HCQ use as pre-exposure prophylaxis to prevent SARS-CoV-2 infection (Table S2; supplementary file). In the meta-analysis, we found no decreased risk of SARS-CoV-2 infection (RR = 0.90; 95% CI 0.46 to 1.77) and no increased risk for any adverse events (RR = 1.36; 95% CI 0.91 to 2.03) for healthcare workers receiving HCQ. The quality of evidence was graded as low for the outcomes of interest (Table 1) (Figures S2 and S3; supplementary file).

**Table 1.**
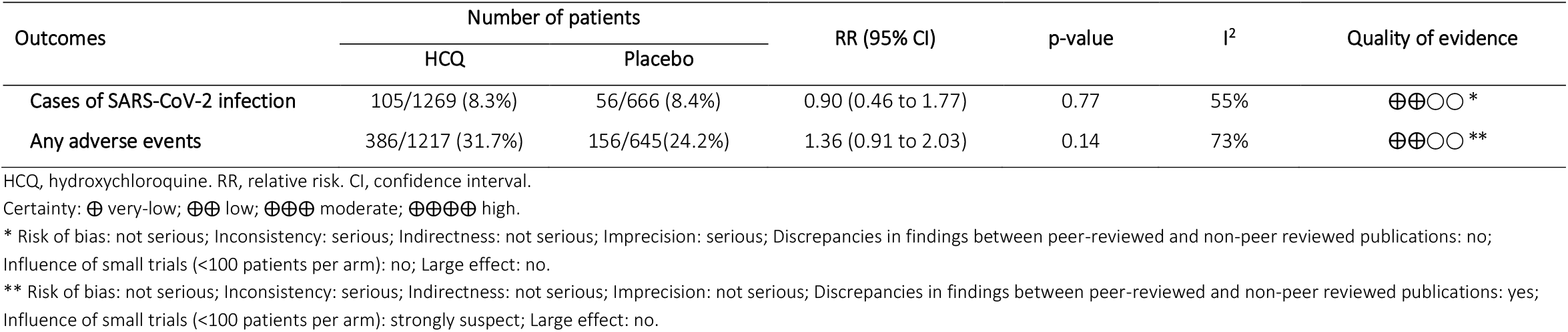
Evidence synthesis on the efficacy and safety of hydroxychloroquine as prophylactic medication pre-exposure to COVID-19.

| Outcomes | Number of patients |  | RR (95% CI) | p-value | I<sup>2</sup> | Quality of evidence |
| --- | --- | --- | --- | --- | --- | --- |
|  | HCQ | Placebo |  |  |  |  |
| **Cases of SARS-CoV-2 infection** | 105/1269 (8.3%) | 56/666 (8.4%) | 0.90 (0.46 to 1.77) | 0.77 | 55% | ⊕⊕○○ * |
| **Any adverse events** | 386/1217 (31.7%) | 156/645(24.2%) | 1.36 (0.91 to 2.03) | 0.14 | 73% | ⊕⊕○○ ** |

HCQ, hydroxychloroquine. RR, relative risk. CI, confidence interval.
Certainty: ⊕ very-low; ⊕⊕ low; ⊕⊕⊕ moderate; ⊕⊕⊕⊕ high.
\* Risk of bias: not serious; Inconsistency: serious; Indirectness: not serious; Imprecision: serious; Discrepancies in findings between peer-reviewed and non-peer reviewed publications: no;
Influence of small trials (<100 patients per arm): no; Large effect: no.
\*\* Risk of bias: not serious; Inconsistency: serious; Indirectness: not serious; Imprecision: not serious; Discrepancies in findings between peer-reviewed and non-peer reviewed publications: yes;
Influence of small trials (<100 patients per arm): strongly suspect; Large effect: no.

This meta-analysis included results of two peer-reviewed studies^33,36^ and two studies published in grey-literature.^34,35^ In the sensitivity analysis, peer-reviewed studies showed no decreased risk of SARS-CoV-2 infection (RR = 0.76; 95% CI 0.52 to 1.10) but an increased risk for any adverse events (RR = 1.60; 95% CI 1.33 to 1.92) among those using HCQ, and the certainty was graded as moderate and high, respectively. No effect on the use of HCQ was found summarizing the results from non-peer reviewed publications and the quality of evidence was classified as very-low (Table S3; supplementary file).

### 3.2 HCQ as post-exposure prophylaxis to prevent SARS-CoV-2 infection

The studies were conducted in the USA and Canada and included adults who had close contact (occupational or household exposure) with a person with known SARS-CoV-2 infection within the prior 96 hours. During trial enrollment in both studies, eligible participants with symptoms consistent with COVID-19 were excluded. All trial protocols were registered on ClinicalTrials.gov, had a parallel design, and were classified as Phase 2/3 or Phase 3. Dosage regimens of HCQ were different between studies and outcomes were assessed within 14 days (Table S4; supplementary file). Studies evaluating HCQ as post-exposure prophylaxis to prevent SARS-CoV-2 infection had a low risk for selection, performance, detection, attrition, reporting, and information bias. However, a high risk for operational bias was detected in both trials (Figure S4; supplementary file).

The two blinded, placebo-controlled RCTs enrolled a total of 1650 individuals, 821 in the HCQ group and 829 in the placebo group. Individual results showed no clinical benefit on HCQ use as post-exposure prophylaxis to prevent SARS-CoV-2 infection (Table S5; supplementary file). In the meta-analysis, we found no decreased risk of SARS-CoV-2 infection (RR = 0.96; 95% CI 0.72 to 1.29) among individuals receiving HCQ, but there was an increased risk for any adverse events within 14 days after trial enrollment (RR = 1.91; 95% CI 1.20 to 3.04). Included studies were peer-reviewed and the quality of evidence was graded as moderate for the outcomes of interest (Table 2) (Figures S5 and S6; supplementary file).

**Table 2.** Evidence synthesis on the efficacy and safety of hydroxychloroquine as prophylactic medication post-exposure to COVID-19.

| Outcomes | Number of patients |  | RR (95% CI) | p-value | I<sup>2</sup> | Quality of evidence |
| --- | --- | --- | --- | --- | --- | --- |
|  | HCQ | Placebo |  |  |  |  |
| Cases of SARS-CoV-2 infection | 102/767 (13.3%) | 103/743 (13.9%) | 0.96 (0.72 to 1.29) | 0.79 | 24% | ⊕⊕⊕○ * |
| Any adverse events | 206/756 (27.3%) | 105/773 (13.6%) | 1.91 (1.20 to 3.04) | < 0.01 | 77% | ⊕⊕⊕○ ** |

HCQ, hydroxychloroquine. RR, relative risk. CI, confidence interval.
Certainty: ⊕ very-low; ⊕⊕ low; ⊕⊕⊕ moderate; ⊕⊕⊕⊕ high.
\* Risk of bias: not serious; Inconsistency: not serious; Indirectness: not serious; Imprecision: serious; Discrepancies in findings between peer-reviewed and non-peer reviewed publications: not applicable; Influence of small trials (<100 patients per arm): not applicable; Large effect: no.
\*\* Risk of bias: not serious; Inconsistency: serious; Indirectness: not serious; Imprecision: not serious; Discrepancies in findings between peer-reviewed and non-peer reviewed publications: not applicable; Influence of small trials (<100 patients per arm): not applicable; Large effect: no.

### 3.3 HCQ as treatment for non-hospitalized patients with COVID-19

The studies were conducted in the USA, Canada and Brazil and included symptomatic, non-hospitalized adults with either laboratory-confirmed SARS-CoV-2 infection or COVID-19-compatible symptoms. All trial protocols were registered on ClinicalTrials.gov, had a parallel design, and were classified as Phase 2/3 or Phase 3. Dosage regimens of HCQ were different between studies and outcomes were assessed between 14 and 90 days (Table S6; supplementary file). All studies evaluating HCQ as treatment of non-hospitalized patients with COVID-19 had a low risk for selection, performance, attrition, and information bias. There was an unclear risk for detection bias in the study by Johnston et al.^28^ and a high risk for reporting bias in the study by Reis et al.^29^ due to the differences in the evaluation time of primary outcomes between trial protocol and full report. All trials were rated as a high risk of operational bias (Figure S7; supplementary file).

The three blinded, placebo-controlled RCTs enrolled a total of 1018 individuals, 497 in the HCQ group and 521 in the placebo group. Individual results showed no clinical benefit on HCQ use as treatment for non-hospitalized patients with COVID-19 (Table S7; supplementary file). In the meta-analysis, we found no decreased risk of hospitalization (RR = 0.64; 95% CI 0.33 to 1.23) and death (RR = 0.62; 95% CI 0.08 to 4.68) and no increased risk of any adverse events (RR = 1.43; 95% CI 0.85 to 2.38) among outpatients with COVID-19 receiving HCQ. Included studies were peer-reviewed and the quality of evidence was graded as moderate for hospitalization and death, and low for any adverse events (Table 3) (Figures S8 – S10; supplementary file).

**Table 3.** Evidence synthesis on the efficacy and safety of hydroxychloroquine as treatment for non-hospitalized patients with COVID-19.

| Outcomes | Number of patients |  | RR (95% CI) | p-value | I<sup>2</sup> | Quality of evidence |
| --- | --- | --- | --- | --- | --- | --- |
|  | HCQ | Placebo |  |  |  |  |
| Hospitalizations | 14/497 (2.8%) | 23/521 (4.4%) | 0.64 (0.33 to 1.23) | 0.18 | 0% | ⊕⊕⊕○ * |
| Deaths | 1/497 (0.2%) | 2/521 (0.4%) | 0.62 (0.08 to 4.68) | 0.64 | 0% | ⊕⊕⊕○ * |
| Any adverse events | 143/490 (29.2%) | 97/514 (18.9%) | 1.43 (0.85 to 2.38) | 0.18 | 72% | ⊕⊕○○ ** |

HCQ, hydroxychloroquine. RR, relative risk. CI, confidence interval.
Certainty: ⊕ very-low; ⊕⊕ low; ⊕⊕⊕ moderate; ⊕⊕⊕⊕ high.
\* / \*\* Risk of bias: not serious; Inconsistency: not serious; Indirectness: not serious; Imprecision: serious; Discrepancies in findings between peer-reviewed and non-peer reviewed publications: not applicable; Influence of small trials (<100 patients per arm): no; Large effect: no.
\*\* Risk of bias: not serious; Inconsistency: serious; Indirectness: not serious; Imprecision: serious; Discrepancies in findings between peer-reviewed and non-peer reviewed publications: not applicable; Influence of small trials (<100 patients per arm): no; Large effect: no.

### 3.4 HCQ as treatment for hospitalized patients with COVID-19

The studies were conducted in the USA, Mexico and France and included non-critically and critically ill patients with COVID-19 requiring hospitalization. All trial protocols were registered on ClinicalTrials.gov, had a parallel design, and were classified as Phase 2 or Phase 3. Dosage regimens of HCQ were similar in most studies and outcomes were assessed up to 14 and 28-30 days (Table S8; supplementary file). Most studies evaluating HCQ as treatment for hospitalized patients with COVID-19 had a low risk for selection, performance, detection, attrition, and information bias. There was potential risk for reporting bias of some outcomes in three studies.^24–26^ All trials were rated as a high risk of operational bias (Figure S11; supplementary file).

The five blinded, placebo-controlled RCTs enrolled a total of 1138 individuals, 572 in the HCQ group and 566 in the placebo group. Individual results showed no clinical benefit on HCQ use as treatment for hospitalized patients with COVID-19 (Table S9; supplementary file). In the meta-analysis, we found no difference in the length of hospital stay between patients treated with HCQ and placebo (MD = 1.20; 95% CI −0.32 to 2.72) (Figure 2). There was no decreased risk of mechanical ventilation ([2 weeks] RR = 0.81; 95% CI 0.49 to 1.34; [4 weeks] RR = 0.97; 95% CI 0.52 to 1.80) and death ([2 weeks] RR = 1.05; 95% CI 0.62 to 1.78; [4 weeks] RR = 0.87; 95% CI 0.67 to 1.13) among hospitalized patients with COVID-19 receiving HCQ. No increased risk of any adverse events was found (RR = 1.07; 95% CI 0.89 to 1.29) (Table 4) (Figures S12 – S16; supplementary file).

**Table 4.** Evidence synthesis on the efficacy and safety of hydroxychloroquine as treatment for hospitalized patients with COVID-19.

| Outcomes | Number of patients |  | RR (95% CI) | p-value | I<sup>2</sup> | Quality of evidence |
| --- | --- | --- | --- | --- | --- | --- |
|  | HCQ | Placebo |  |  |  |  |
| **Mechanical ventilation** |  |  |  |  |  |  |
| 2 weeks | 26/433 (6.0%) | 31/421 (7.4%) | 0.81 (0.49 to 1.34) | 0.42 | 0% | ⊕⊕⊕○ * |
| 4 weeks | 19/433 (4.4%) | 19/421 (4.5%) | 0.97 (0.52 to 1.80) | 0.91 | 0% | ⊕⊕⊕○ * |
| **Death** |  |  |  |  |  |  |
| 2 weeks | 27/433 (6.2%) | 25/421 (5.9%) | 1.05 (0.62 to 1.78) | 0.86 | 0% | ⊕⊕⊕○ * |
| 4 weeks | 80/572 (14.0%) | 92/566 (16.3%) | 0.87 (0.67 to 1.13) | 0.30 | 0% | ⊕⊕⊕○ * |
| **Any adverse events** | 122/433 (28.2%) | 108/418 (25.8%) | 1.07 (0.89 to 1.29) | 0.45 | 0% | ⊕⊕⊕○ * |

HCQ, hydroxychloroquine. RR, relative risk. CI, confidence interval.
Certainty: ⊕ very-low; ⊕⊕ low; ⊕⊕⊕ moderate; ⊕⊕⊕⊕ high.
\* Risk of bias: not serious; Inconsistency: not serious; Indirectness: not serious; Imprecision: serious; Discrepancies in findings between peer-reviewed and non-peer reviewed publications: no / not applicable; Influence of small trials (<100 patients per arm): no; Large effect: no.

**Figure 2.**
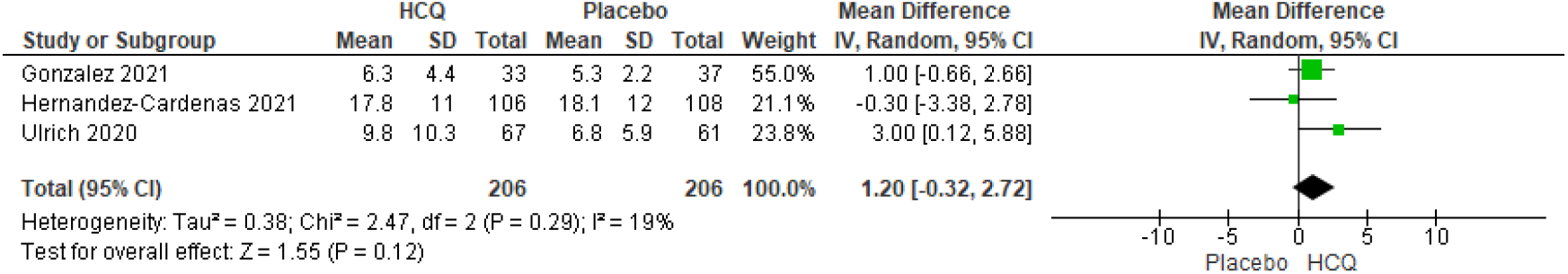
Forest plot showing differences in length of hospital stay between patients treated with HCQ and placebo.

All trials evaluating the need for mechanical ventilation and any adverse events were peer-reviewed as well as trials reporting data on deaths during two weeks of follow-up. Of the five trials reporting data on deaths during four weeks of follow-up, three were peer-reviewed^23,24,27^ and two were non-peer reviewed publications.^25,26^ The quality of evidence was graded as moderate for the outcomes of interest (Table 4). In the sensitivity analysis for mortality during four weeks of follow-up, we found no decreased risk for death from peer reviewed (RR = 0.88; 95% CI 0.58 to 1.33) and non-peer reviewed (RR = 0.80; 95% CI 0.42 to 1.55) publications, and the quality of evidence was also classified as moderate (Table S10; supplementary file).

### 3.5 Risk of any and serious adverse events, QTc interval >500ms and other cardiac manifestations, gastrointestinal symptoms, headache, rash, and neurologic reactions for individuals using HCQ

We found an increased risk for any adverse events (RR = 1.38; 95% CI 1.12 to 1.71) and gastrointestinal symptoms (RR = 2.45; 95% CI 1.77 to 3.39) among individuals treated with HCQ. However, there was no increased risk for serious adverse events (RR = 1.07; 95% CI 0.69 to 1.67), QTc >500ms (RR = 2.13; 95% CI 0.96 to 4.71) and other cardiac manifestations (RR = 1.52; 95% 0.60 to 3.83), headache (RR = 0.93; 95% CI 0.55 to 1.56), skin rash (RR = 1.46; 95% CI 0.89 to 2.29), and neurologic reactions (RR = 1.20; 95% CI 0.80 to 1.78) (Table 5) (Figures S17 – S24; supplementary file).

**Table 5.**
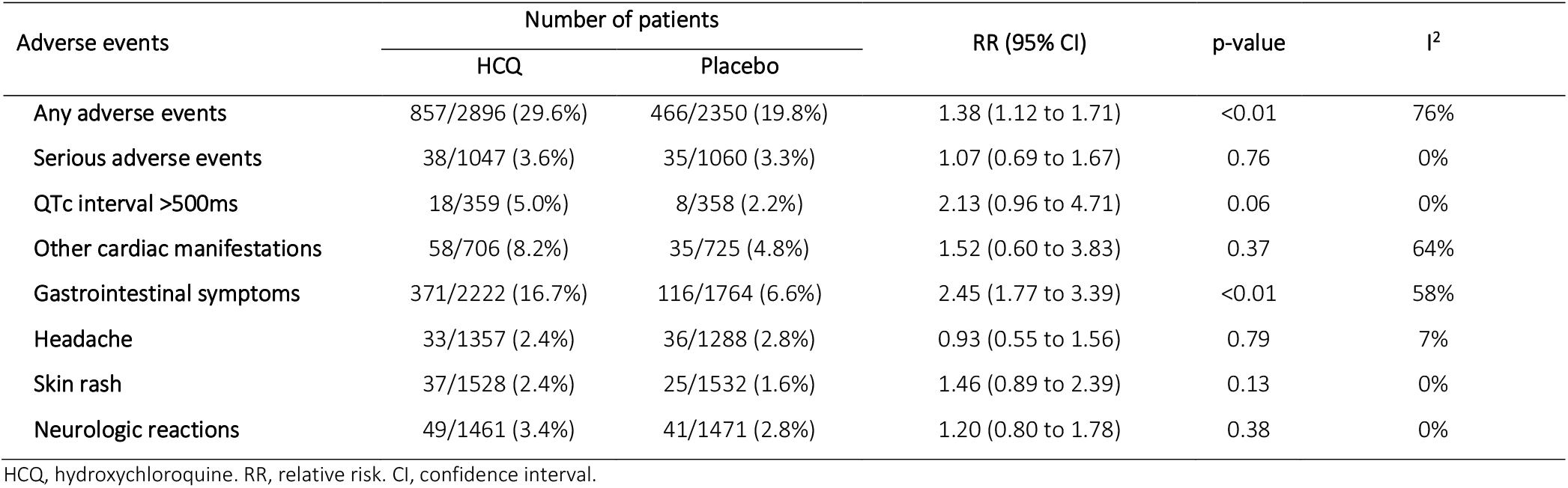
Evidence synthesis on the safety of hydroxychloroquine for COVID-19 based on the results of blinded, placebo-controlled, randomized clinical trials.

| Adverse events | Number of patients |  | RR (95% CI) | p-value | I<sup>2</sup> |
| --- | --- | --- | --- | --- | --- |
|  | HCQ | Placebo |  |  |  |
| Any adverse events | 857/2896 (29.6%) | 466/2350 (19.8%) | 1.38 (1.12 to 1.71) | <0.01 | 76% |
| Serious adverse events | 38/1047 (3.6%) | 35/1060 (3.3%) | 1.07 (0.69 to 1.67) | 0.76 | 0% |
| QTc interval >500ms | 18/359 (5.0%) | 8/358 (2.2%) | 2.13 (0.96 to 4.71) | 0.06 | 0% |
| Other cardiac manifestations | 58/706 (8.2%) | 35/725 (4.8%) | 1.52 (0.60 to 3.83) | 0.37 | 64% |
| Gastrointestinal symptoms | 371/2222 (16.7%) | 116/1764 (6.6%) | 2.45 (1.77 to 3.39) | <0.01 | 58% |
| Headache | 33/1357 (2.4%) | 36/1288 (2.8%) | 0.93 (0.55 to 1.56) | 0.79 | 7% |
| Skin rash | 37/1528 (2.4%) | 25/1532 (1.6%) | 1.46 (0.89 to 2.39) | 0.13 | 0% |
| Neurologic reactions | 49/1461 (3.4%) | 41/1471 (2.8%) | 1.20 (0.80 to 1.78) | 0.38 | 0% |

HCQ, hydroxychloroquine. RR, relative risk. CI, confidence interval.

## 4 DISCUSSION

There is still no optimal approach toward COVID-19 management and several repurposing drugs including remdesivir, ivermectin, colchicine, favipiravir, lopinavir-ritonavir, ribavirin, interferon, CQ and HCQ have been tested for prevention and treatment of disease. Despite drug repurposing played a critical role in the identification of rapidly available therapeutic solutions against SARS-CoV-2 infection^37^, to date only remdesivir and tocilizumab were approved by the US Food and Drug Administration (FDA) and other healthy agencies for the treatment of hospitalized patients with COVID-19. The evidence for repurposing HCQ is based on decreased viral replication *in vitro* but results from clinical data are contrasting. In this systematic review and meta-analysis, we synthesized the available evidence on the efficacy and safety of HCQ in prevention and treatment of COVID-19 based on the results of blinded, placebo-controlled RCTs. Our findings confirm the ineffectiveness of HCQ against COVID-19 in the current state of the art as established by best practices.

HCQ, a 4-aminoquinoline compound used as an antimalarial drug, was reported to be effective in inhibiting SARS-CoV-2 infection *in vitro* due to the intracellular pH change and interference with the glycosylation of ACE2 receptor and the spike protein leading to block the virus entry into target cells.^1^ Despite promising preclinical results, HCQ has not been shown effective as pre- and post-exposure prophylaxis to COVID-19. The National Institutes of Health (NIH)^38^, WHO ^11^ and the European Medical Agency (EMA)^39^ have not recommend the use of any drug as prophylaxis against SARS-CoV-2 infection, except in clinical trials. The use of general prevention measures such as mask wearing, hand hygiene, and physical distancing remain indicated and are effective nonpharmacologic interventions to reduce the spread of this infection.^40,41^ In addition, it has been shown that vaccines currently available are 50%-95% effective in preventing COVID-19.^42–47^ Mass vaccination is the most cost-effective measure for controlling and preventing SARS-CoV-2 infection.

Currently, there is a consensus that symptomatic cases of COVID-19 require supportive care with medical evaluation, stratification of risk factors for worse clinical outcomes^48–50^, clinical monitoring of symptoms, and complementary exams if indicated. Measures to reduce the risk of SARS-CoV-2 transmission as patient isolation should also be guided. In outpatients, symptomatic treatment includes analgesics and antipyretics. For patients with dyspnea, prone positioning and respiratory physiotherapy may be indicated.^51,52^ Regular fluid intake should also be advised to avoid dehydration, and walking should be encouraged according to the patient’s tolerance. SARS-CoV-2 specific monoclonal antibodies (bamlanivimab, bamlanivimab with etesevimab, and casirivimab with imdevimab) have been considered in the treatment of outpatients with mild to moderate COVID-19, in the presence of high-risk criteria.^53–58^ In clinical trials, HCQ has not been associated with a reduction in the prevalence, severity, or duration of COVID-19 symptoms^28–30^, and there is no decreased risk of hospitalization for outpatients with SARS-CoV-2 infection treated with the drug. One of the hypotheses raised for the lack of response of HCQ in the clinical trials is that patients would have achieved minimal concentrations lower than the target concentration identified from *in vitro* assays. However, in at least one trial, HCQ dose was designed to achieve and maintain drug concentration above half the estimated maximum effective concentration (EC50) for SARS-CoV-2.^30^

Patients with persistent dyspnea, chest pain, hypoxia with a measured oxygen saturation below 94%, altered mental status, or persistent fever should be evaluated for the need of hospital admission. In hospitalized patients, treatment includes supplemental oxygen and prone position when necessary, and adequate management of pulmonary ventilation.^59–62^ Extracorporeal membrane oxygenation has been indicated in severe refractory cases.^63–66^ For some patients, SARS-CoV-2 infection may lead to a massive release of proinflammatory cytokines and hypercoagulable state increasing the risk for thromboembolic events.^67–69^ In clinical practice, the decision to prescribe low molecular weight heparin (LMWH) or unfractionated heparin has been made on a case-by-case basis.^70^ Despite real world evidence recommending the use of prophylactic anticoagulation for patients with COVID-19 on hospital admission^71^, recent open-label RCTs^72,73^ showed that prophylactic or therapeutic anticoagulation did not result in clinical improvement for hospitalized patients with COVID-19, except in the context of diagnosing a thromboembolic event.

In hospitalized patients with COVID-19, dexamethasone was shown to reduce mortality in patients who required supplemental oxygen.^74^ Remdesivir, an FDA-approved intravenous antiviral drug, has been associated in double-blinded, placebo-controlled RCTs with a faster time to clinical improvement in severe cases of COVID-19, but results on mortality are contrasting.^75,76^ Recently, there is emerging evidence on the efficacy of tocilizumab, an interleukin-6 inhibitor, on clinical outcomes in critical COVID-19 patients with systemic inflammation and rapid respiratory deterioration.^77,78^ In hospital setting, our results found no difference in the length of stay and no decreased risk for mechanical ventilation and deaths among patients treated with HCQ compared to placebo.

In addition to not having found clinical benefits of HCQ to prevent or treat COVID-19, our results demonstrated an increased risk for any adverse events and gastrointestinal symptoms among those using HCQ. Although a higher proportion of individuals treated with HCQ had QTc interval > 500ms compared to placebo (5.0% vs. 2.2%), there is no statistical difference between groups and no cases of arrhythmia or death associated to HCQ were reported. However, we must consider the fact that patients at higher risk of cardiotoxicity were excluded from these studies and the indiscriminate use of these drugs should be avoided.

Out results had limitations and included trials with a high risk for operational bias. In addition, we found an important influence of non-peer reviewed studies in the quality of evidence for some outcomes of interest. However, the certainty of the results on the lack of clinical benefit for HCQ was rated as moderate and we believe that the true effect is probably close to the estimated effect. Finally, we lack in analyzing potential adverse events dose-response relationships in patients treated with HCQ.

## 5 CONCLUSION

Available evidence based on the results of blinded, placebo-controlled RCTs showed no clinical benefits of HCQ as pre-and post-exposure prophylaxis and treatment of non-hospitalized and hospitalized patients with COVID-19.

## Data Availability

The data sets used and/or analyzed during the current study are available from the corresponding author on reasonable request.

## ACKNOWLEDGMENTS

To all health professionals who are facing the COVID-19 pandemic.

