## Supplementary file for "Efficacy and safety of hydroxychloroquine as pre-and post-exposure prophylaxis and treatment of COVID-19. Systematic review and meta-analysis of blinded, placebo-controlled, randomized clinical trials"

### Search strategy

#### PubMed:

(Chloroquine OR Chlorochin OR Arequin OR Aralen OR Arechine OR Nivaquine OR Chingamin OR Hydroxychloroquine OR Hydroxychlorochin OR Oxychlorochin OR Plaquenil) AND (COVID-19 OR “2019-nCoV Infection” OR “Coronavirus Disease-19” OR “2019-nCoV Disease” OR SARS-CoV-2) AND (randomized controlled trial[Publication Type] OR (randomized[Title/Abstract] AND controlled[Title/Abstract] AND trial[Title/Abstract]))

#### Embase:

(Chloroquine OR Chlorochin OR Arequin OR Aralen OR Arechine OR Nivaquine OR Chingamin OR Hydroxychloroquine OR Hydroxychlorochin OR Oxychlorochin OR Plaquenil) AND (COVID-19 OR “2019-nCoV Infection” OR “Coronavirus Disease-19” OR “2019-nCoV Disease” OR SARS-CoV-2) AND ('crossover procedure':de OR 'double-blind procedure':de OR 'randomized controlled trial':de OR 'single-blind procedure':de OR (random* OR factorial* OR crossover* OR cross NEXT/1 over* OR placebo* OR doubl* NEAR/1 blind* OR singl* NEAR/1 blind* OR assign* OR allocat* OR volunteer*):de,ab,ti)

#### Web of Science:

(Chloroquine OR Chlorochin OR Arequin OR Aralen OR Arechine OR Nivaquine OR Chingamin OR Hydroxychloroquine OR Hydroxychlorochin OR Oxychlorochin OR Plaquenil) AND (COVID-19 OR “2019-nCoV Infection” OR “Coronavirus Disease-19” OR “2019-nCoV Disease” OR SARS-CoV-2) AND (TS= clinical trial* OR TS=research design OR TS=comparative stud* OR TS=evaluation stud* OR TS=controlled trial* OR TS=follow-up stud* OR TS=prospective stud* OR TS=random* OR TS=placebo* OR TS=(single blind*) OR TS=(double blind*))

#### Lilacs:

(Chloroquine OR Chlorochin OR Arequin OR Aralen OR Arechine OR Nivaquine OR Chingamin OR Hydroxychloroquine OR Hydroxychlorochin OR Oxychlorochin OR Plaquenil) AND (COVID-19 OR “2019-nCoV Infection” OR “Coronavirus Disease-19” OR “2019-nCoV Disease” OR SARS-CoV-2) AND (“clinical trial” OR placebo OR “controlled trial” OR “double blind”)

#### medRxiv:

(Chloroquine OR Hydroxychloroquine) AND (COVID-19 OR SARS-CoV-2) AND (“clinical trial” OR placebo OR “controlled trial”)

### Hydroxychloroquine as pre-exposure prophylaxis to prevent SARS-CoV-2 infection

Table S1. Blinded, placebo-controlled, randomized clinical trials evaluating the efficacy and safety of hydroxychloroquine as prophylactic medication pre-exposure to COVID-19.

| Author | Study | Country | Protocol and Phase | Population | Intervention | Control | Outcomes of interest (Time of evaluation) |
|---|---|---|---|---|---|---|---|
| Abella 2021 | Efficacy and Safety of Hydroxychloroquine vs Placebo for Pre-exposure SARS-cov-2 Prophylaxis Among Health Care Workers: A Randomized Clinical Trial | USA | NCT04329923 (Phase 2) | Healthcare workers at high-risk for SARS-CoV-2 infection | HCQ 600mg/day for 8 weeks (n = 66) | Placebo (n = 66) | Incidence of SARS-CoV-2 infection and adverse events (8 weeks) |
| Rojas-Serrano 2021 | Hydroxychloroquine for Prophylaxis Of COVID-19 in Health Workers: A Randomized Clinical Trial | Mexico | NCT04318015 (Phase 3) | Healthcare workers at high-risk for SARS-CoV-2 infection | HCQ 200mg/day for 8 weeks (n = 62) | Placebo (n = 65) | Incidence of SARS-CoV-2 infection and adverse events (8 weeks) |
| Syed 2021 | Pre-Exposure Prophylaxis with Various Doses of Hydroxychloroquine among high-risk COVID 19 Healthcare Personnel: CHEER randomized controlled trial | Pakistan | NCT04359537 (Phase 2) | Healthcare workers at high-risk for SARS-CoV-2 infection | A: HCQ 400mg, 2x/day in the first day + 400mg 1x/week for 12 weeks<br>B: HCQ 400mg, once every 3 weeks<br>C: HCQ 200mg, once every 3 weeks<br>(A: n = 48)<br>(B: n = 51)<br>(C: n = 55) | Placebo (n = 46) | Incidence of SARS-CoV-2 infection and adverse events (12 weeks) |
| Rajasingham 2020 | Hydroxychloroquine as pre-exposure prophylaxis for COVID-19 in healthcare workers: a randomized trial | USA and Canada | NCT04328467 (Phase 3) | Healthcare workers at high-risk for SARS-CoV-2 infection | A: HCQ 400mg loading dose + 400mg 1x/week for 12 weeks<br>B: HCQ 400mg loading dose + 400mg 2x/weeks for 12 weeks<br>(A: n = 494)<br>(B: n = 495) | Placebo (n = 494) | Incidence of SARS-CoV-2 infection and adverse events (12 weeks) |

HCQ, hydroxychloroquine.

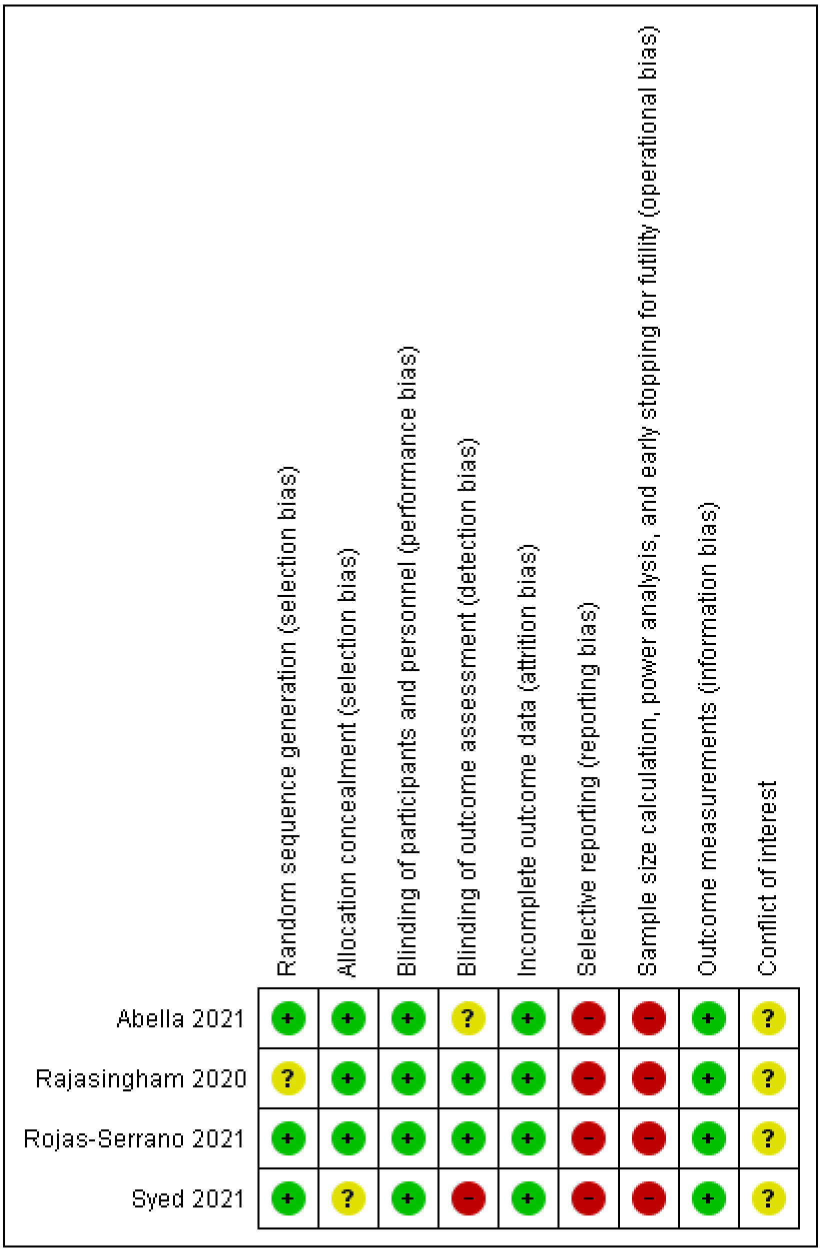


Fig. S1. Risk of bias assessment on the use of hydroxychloroquine as prophylactic medication pre-exposure to COVID-19.

Table S2. Individual results of studies evaluating the efficacy and safety of hydroxychloroquine as prophylactic medication pre-exposure to COVID-19.

| Author | Cases of COVID-19 | Any adverse events | Conclusion |
|---|---|---|---|
| Abella 2021 | HCQ: 4/64 (6.3%)<br>Placebo: 4/61 (6.6%) | HCQ: 29/65 (44.6%)<br>Placebo: 17/65 (26.2%) | “There was no clinical benefit of hydroxychloroquine administered daily for 8 weeks as pre-exposure prophylaxis in hospital-based HCWs exposed to patients with COVID-19.” |
| Rojas-Serrano 2021 | HCQ: 1/62 (1.6%)<br>Placebo: 6/65 (9.2%) | HCQ: 32/62 (51.6%)<br>Placebo: 38/65 (58.5%) | “No beneficial effect or significant harm could be demonstrated in our randomized controlled trial including 127 participants, using relatively low doses of HCQ compared with placebo in health personnel.” |
| Syed 2021 | A: HCQ 400mg, 1x/week: 15/48 (31.3%)<br>B: 400mg, once every 3 weeks: 19/51 (37.3%)<br>C: 200mg, once every 3 weeks: 8/55 (14.5%)<br>Placebo: 7/46 (15.2%) | A: HCQ 400mg, 1x/week: 5/48 (10.4%)<br>B: 400mg, once every 3 weeks: 1/51 (2.0%)<br>C: 200mg, once every 3 weeks: 3/55 (5.5%)<br>Placebo: 1/46 (2.2%) | “There was no significant reduction in the SARS-CoV-2 transmission with PrEP administration of hydroxychloroquine among the enrolled healthcare personnel.” |
| Rajasingham 2020 | A: HCQ 400mg, 1x/week: 29/494 (5.9%)<br>B: HCQ 400mg, 2x/week: 29/495 (5.9%)<br>Placebo: 39/494 (7.9%) | A: HCQ 400mg, 1x/week: 148/473 (31.3%)<br>B: HCQ 400mg, 2x/week: 168/463 (36.3%)<br>Placebo: 100/469 (21.3%) | “Pre-exposure prophylaxis with hydroxychloroquine once or twice weekly did not significantly reduce laboratory-confirmed COVID-19 or COVID-19–compatible illness among healthcare workers.” |

HCQ, hydroxychloroquine.

| | HCQ | | Placebo | | | Risk Ratio |
|---|---|---|---|---|---|---|
| **Study or Subgroup** | **Events** | **Total** | **Events** | **Total** | **Weight** | **M-H, Random, 95% CI** |
| Abella 2021 | 4 | 64 | 4 | 61 | 16.7% | 0.95 [0.25, 3.64] |
| Rajasingham 2020 | 58 | 989 | 39 | 494 | 43.1% | 0.74 [0.50, 1.10] |
| Rojas-Serrano 2021 | 1 | 62 | 6 | 65 | 8.6% | 0.17 [0.02, 1.41] |
| Syed 2021 | 42 | 154 | 7 | 46 | 31.6% | 1.79 [0.86, 3.72] |
| **Total (95% CI)** | | **1269** | | **666** | **100.0%** | **0.90 [0.46, 1.77]** |
| Total events | 105 | | 56 | | | |

Heterogeneity: Tau² = 0.23; Chi² = 6.69, df = 3 (P = 0.08); I² = 55%
Test for overall effect: Z = 0.30 (P = 0.77)

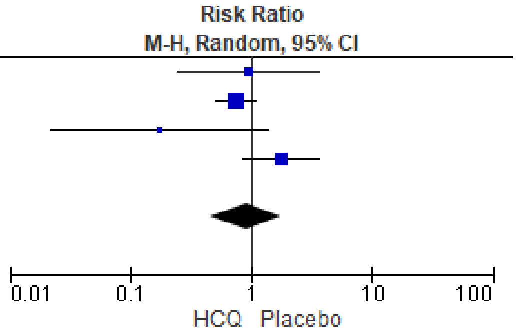


Fig. S2. Forest plot showing the risk of SARS-CoV-2 infection for individuals using hydroxychloroquine as prophylactic medication pre-exposure to COVID-19.

| | HCQ | | Placebo | | | Risk Ratio |
|---|---|---|---|---|---|---|
| **Study or Subgroup** | **Events** | **Total** | **Events** | **Total** | **Weight** | **M-H, Random, 95% CI** |
| Abella 2021 | 29 | 65 | 17 | 65 | 25.7% | 1.71 [1.04, 2.78] |
| Rajasingham 2020 | 316 | 936 | 100 | 469 | 37.7% | 1.58 [1.30, 1.93] |
| Rojas-Serrano 2021 | 32 | 62 | 38 | 65 | 33.0% | 0.88 [0.64, 1.21] |
| Syed 2021 | 9 | 154 | 1 | 46 | 3.6% | 2.69 [0.35, 20.67] |
| **Total (95% CI)** | | **1217** | | **645** | **100.0%** | **1.36 [0.91, 2.03]** |
| Total events | 386 | | 156 | | | |

Heterogeneity: Tau² = 0.10; Chi² = 11.17, df = 3 (P = 0.01); I² = 73%
Test for overall effect: Z = 1.48 (P = 0.14)

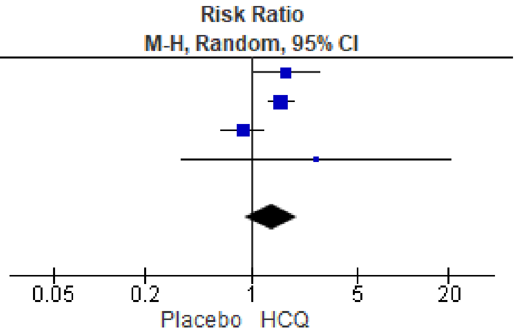


Fig. S3. Forest plot showing the risk of any adverse events for individuals using hydroxychloroquine as prophylactic medication pre-exposure to COVID-19.

Table S3. Sensitivity analysis of studies evaluating the efficacy and safety of hydroxychloroquine as prophylactic medication pre-exposure to COVID-19.

| Outcomes | RR (95% CI) | $I^2$ | Risk of bias | Inconsistency | Indirectness | Imprecision | Influence of small trials | Large effect | Quality of evidence |
|---|---|---|---|---|---|---|---|---|---|
| SARS-CoV-2 infection | | | | | | | | | |
| Peer-reviewed studies | 0.76 (0.52 to 1.10) | 0% | Not serious | Not serious | Not serious | Serious | Not serious | No | ⊕⊕⊕◯ |
| Non-peer reviewed studies | 0.69 (0.07 to 6.73) | 77% | Serious | Serious | Not serious | Serious | Strongly suspect | No | ⊕◯◯◯ |
| Any adverse events | | | | | | | | | |
| Peer-reviewed studies | 1.60 (1.33 to 1.92) | 0% | Serious | Not serious | Not serious | Not serious | Not serious | No | ⊕⊕⊕⊕ |
| Non-peer reviewed studies | 1.00 (0.49 to 2.05) | 18% | Serious | Serious | Not serious | Serious | Not serious | No | ⊕◯◯◯ |

Certainty: ⊕ very-low; ⊕⊕ low; ⊕⊕⊕ moderate; ⊕⊕⊕⊕ high.

Table S4. Blinded, placebo-controlled, randomized clinical trials evaluating the efficacy and safety of hydroxychloroquine as prophylactic medication post-exposure to COVID-19.

| Author | Study | Country | Protocol and Phase | Population | Intervention | Control | Outcomes of interest (Time of evaluation) |
|---|---|---|---|---|---|---|---|
| Barnabas 2020 | Hydroxychloroquine as Postexposure Prophylaxis to Prevent Severe Acute Respiratory Syndrome Coronavirus 2 Infection: A Randomized Trial | USA | NCT04328961 (Phase 2/3) | Asymptomatic adults who had close contact (occupational or household exposure) with a person with recent known SARS-CoV-2 infection | HCQ 400mg/day per 3 days + 200mg/day for 11 days (n = 407) | Placebo (n = 422) | Incidence of SARS-CoV-2 infection and adverse events (14 days) |
| Boulware 2020 | A Randomized Trial of Hydroxychloroquine as Postexposure Prophylaxis for Covid-19 | USA and Canada | NCT04308668 (Phase 3) | Asymptomatic adults who had close contact (occupational or household exposure) with a person with recent known SARS-CoV-2 infection | HCQ 800mg loading dose followed by 600mg in 6 to 8h + 600mg/day for 4 days (n = 414) | Placebo (n = 407) | Incidence of SARS-CoV-2 infection and adverse events (14 days) |

HCQ, hydroxychloroquine.

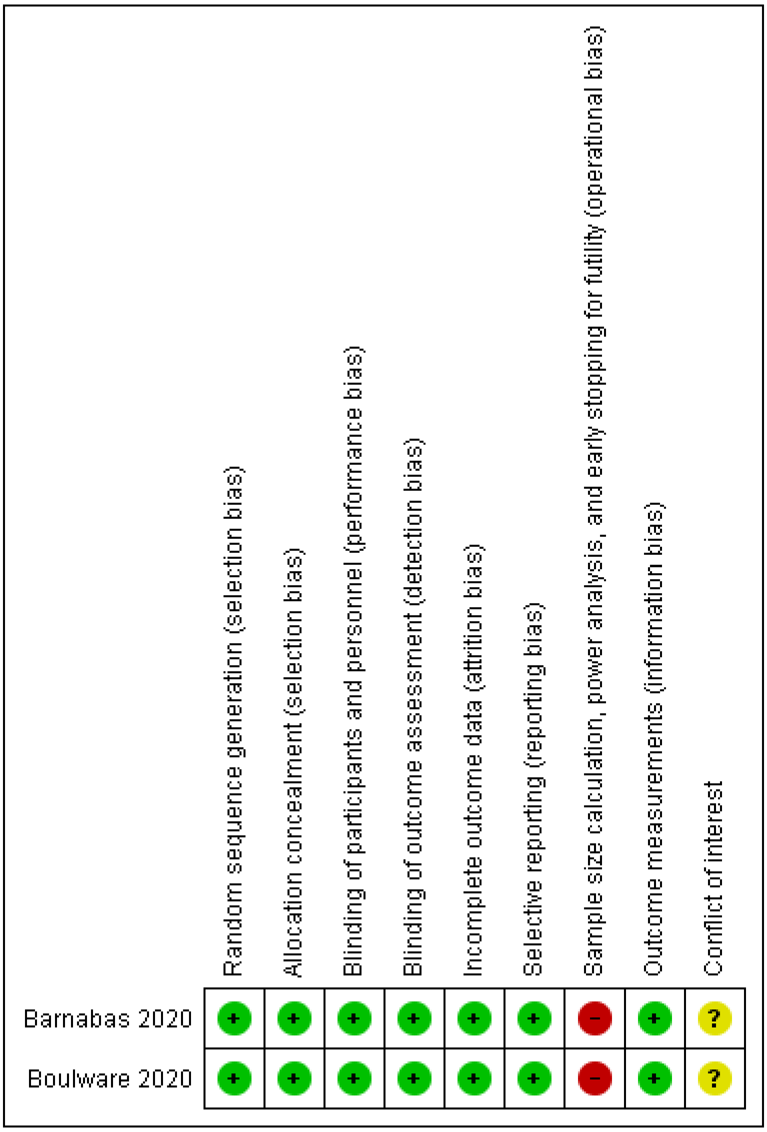


Fig. S4. Risk of bias assessment on the use of hydroxychloroquine as prophylactic medication post-exposure to COVID-19.

Table S5. Individual results of studies evaluating the efficacy and safety of hydroxychloroquine as prophylactic medication post-exposure to COVID-19.

| Author | Cases of COVID-19 | Any adverse events | Conclusion |
|---|---|---|---|
| Barnabas 2020 | HCQ: 53/353 (15.0%)<br>Placebo: 45/336 (13.4%) | HCQ: 66/407 (16.2%)<br>Placebo: 46/422 (10.9%) | "This rigorous randomized controlled trial among persons with recent exposure excluded a clinically meaningful effect of hydroxychloroquine as postexposure prophylaxis to prevent SARS-CoV-2 infection." |
| Boulware 2020 | HCQ: 49/414 (11.8%)<br>Placebo: 58/407 (14.3%) | HCQ: 140/349 (40.1%)<br>Placebo: 59/351 (16.8%) | "This randomized trial did not demonstrate a significant benefit of hydroxychloroquine as postexposure prophylaxis for Covid-19." |

HCQ, hydroxychloroquine.

| Study or Subgroup | HCQ Events | HCQ Total | Placebo Events | Placebo Total | Weight | Risk Ratio M-H, Random, 95% CI |
|---|---|---|---|---|---|---|
| Barnabas 2020 | 53 | 353 | 45 | 336 | 48.6% | 1.12 [0.78, 1.62] |
| Boulware 2020 | 49 | 414 | 58 | 407 | 51.4% | 0.83 [0.58, 1.18] |
| **Total (95% CI)** | | **767** | | **743** | **100.0%** | **0.96 [0.72, 1.29]** |
| Total events | 102 | | 103 | | | |

Heterogeneity: Tau² = 0.01; Chi² = 1.32, df = 1 (P = 0.25); I² = 24%
Test for overall effect: Z = 0.27 (P = 0.79)

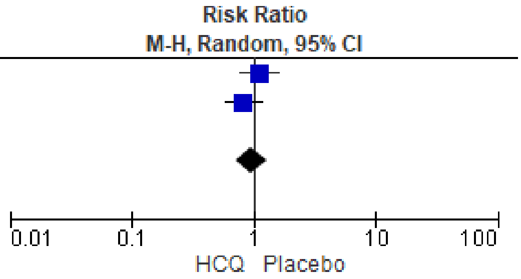


Fig. S5. Forest plot showing the risk of SARS-CoV-2 infection for individuals using hydroxychloroquine as prophylactic medication post-exposure to COVID-19.

| Study or Subgroup | HCQ Events | HCQ Total | Placebo Events | Placebo Total | Weight | Risk Ratio M-H, Random, 95% CI |
|---|---|---|---|---|---|---|
| Barnabas 2020 | 66 | 407 | 46 | 422 | 46.9% | 1.49 [1.05, 2.11] |
| Boulware 2020 | 140 | 349 | 59 | 351 | 53.1% | 2.39 [1.83, 3.11] |
| **Total (95% CI)** | | **756** | | **773** | **100.0%** | **1.91 [1.20, 3.04]** |
| Total events | 206 | | 105 | | | |

Heterogeneity: Tau² = 0.09; Chi² = 4.43, df = 1 (P = 0.04); I² = 77%
Test for overall effect: Z = 2.75 (P = 0.006)

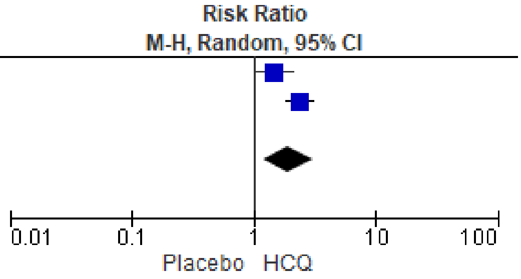


Fig. S6. Forest plot showing the risk of any adverse events for individuals using hydroxychloroquine as prophylactic medication post-exposure to COVID-19.

Table S6. Blinded, placebo-controlled, randomized clinical trials evaluating the efficacy and safety of hydroxychloroquine as treatment for non-hospitalized patients with COVID-19.

| Author | Study | Country | Protocol and Phase | Population | Intervention | Control | Outcomes of interest (Time of evaluation) |
|---|---|---|---|---|---|---|---|
| Skipper 2020 | Hydroxychloroquine in Nonhospitalized Adults with Early COVID-19: A Randomized Trial | USA and Canada | NCT04308668 (Phase 3) | Symptomatic, non-hospitalized patients with RT-PCR diagnosis of COVID-19 or compatible symptoms after a high-risk exposure to a person with known COVID-19<br>• HCQ group: median age 41y (range 33-49). Sex: 42.0% male. Medical conditions: hypertension 10.8%, diabetes 3.8%.<br>• Placebo: median age 39y (range 31-50). Sex: 45.5% male. Medical conditions: hypertension 10.9%, diabetes 3.3%. | HCQ 800mg loading dose followed by 600mg in 6 to 8h + 600mg/day for 4 days (n = 212) | Placebo (n = 211) | Hospitalization, death, and adverse events (14 days) |
| Reis 2021 | Effect of Early Treatment with Hydroxychloroquine or Lopinavir and Ritonavir on Risk of Hospitalization Among Patients With COVID-19. The TOGETHER Randomized Clinical Trial | Brazil | NCT04403100 (Phase 3) | Symptomatic, non-hospitalized patients with RT-PCR diagnosis of COVID-19 or a clinical condition compatible with COVID-19 and respiratory symptoms<br>• HCQ group: median age 53y (range 18-81). Sex: 43.0% male. Medical conditions: hypertension 47.2%, diabetes: 19.2%, heart disease: 2.8%.<br>• Placebo: median age 53y (range 18-80). Sex: 46.7% male. Medical conditions: hypertension 48.0%, diabetes: 21.1%, heart disease: 3.5% | HCQ 800mg loading dose + 400mg/day for 9 days (n = 214) | Placebo (n = 227) | Hospitalization, death, and adverse events (90 days) |
| Johnston 2021 | Hydroxychloroquine with or without azithromycin for treatment of early SARS-CoV-2 infection among high-risk outpatient adults: A randomized clinical trial | USA | NCT04354428 (Phase 2/3) | Symptomatic (> 92%), non-hospitalized patients with RT-PCR diagnosis of COVID-19<br>• HCQ group: median age 36y (range 19-78). Sex: 45.1% male. Medical conditions: hypertension 11.3%, diabetes 7.0%.<br>• Placebo: median age 38y (range 18-70). Sex: 45.8% male. Medical conditions: hypertension 8.4%, diabetes 6.0%. | HCQ 400mg, 2x/day in the first day + 200mg, 2x/day for 9 days (n = 71) | Placebo (n = 83) | Hospitalization, death, and adverse events (28 days) |

HCQ, hydroxychloroquine.

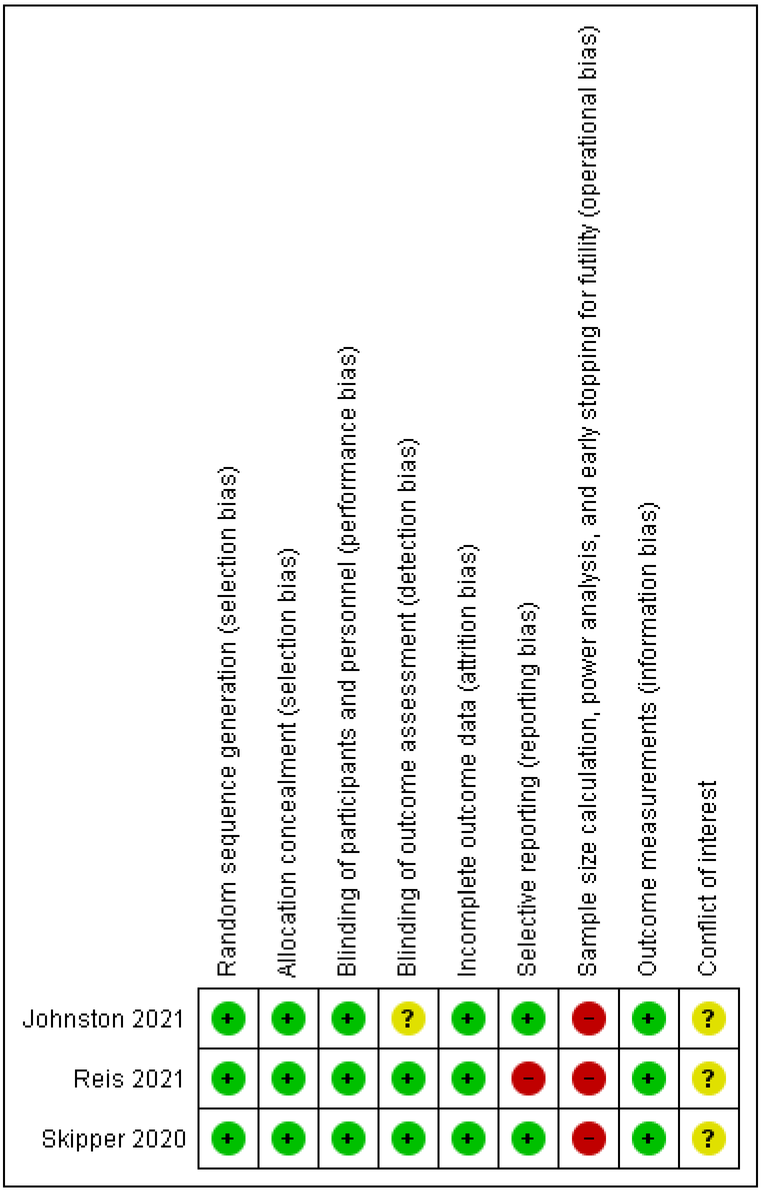


Fig. S7. Risk of bias assessment on the use of hydroxychloroquine as treatment for non-hospitalized patients with COVID-19.

Table S7. Individual results of studies evaluating the efficacy and safety of hydroxychloroquine as treatment for non-hospitalized patients with COVID-19.

| Author | Need of hospitalization | Death | Any adverse events | Conclusion |
|---|---|---|---|---|
| Skipper 2020 | HCQ: 4/212 (1.9%)<br>Placebo: 8/211 (3.8%) | HCQ: 1/212 (0.5%)<br>Placebo: 1/211 (0.5%) | HCQ: 92/212 (43.4%)<br>Placebo: 46/211 (21.8%) | "Hydroxychloroquine did not substantially reduce symptom severity in outpatients with early, mild COVID-19." |
| Reis 2021 | HCQ: 8/214 (3.7%)<br>Placebo: 11/227 (4.9%) | HCQ: 0/214<br>Placebo: 1/227 (0.4%) | HCQ: 46/207 (22.2%)<br>Placebo: 46/220 (20.9%) | "In this randomized clinical trial, neither hydroxychloroquine nor lopinavir-ritonavir showed any significant benefit for decreasing COVID-19–associated hospitalization or other secondary clinical outcomes." |
| Johnston 2021 | HCQ: 2/71 (2.8%)<br>Placebo: 4/83 (4.8%) | HCQ: 0/71<br>Placebo: 0/83 | HCQ: 5/71 (7.0%)<br>Placebo: 5/83 (6.0%) | "HCQ and HCQ/AZ are not effective therapies for outpatient treatment of SARV-CoV-2 infection." |

HCQ, hydroxychloroquine.

| Study or Subgroup | HCQ Events | HCQ Total | Placebo Events | Placebo Total | Weight | Risk Ratio M-H, Fixed, 95% CI |
|---|---|---|---|---|---|---|
| Johnston 2021 | 2 | 71 | 4 | 83 | 16.5% | 0.58 [0.11, 3.10] |
| Reis 2021 | 8 | 214 | 11 | 227 | 47.7% | 0.77 [0.32, 1.88] |
| Skipper 2020 | 4 | 212 | 8 | 211 | 35.8% | 0.50 [0.15, 1.63] |
| **Total (95% CI)** | | **497** | | **521** | **100.0%** | **0.64 [0.33, 1.23]** |
| Total events | 14 | | 23 | | | |

Heterogeneity: Chi² = 0.35, df = 2 (P = 0.84); I² = 0%
Test for overall effect: Z = 1.33 (P = 0.18)

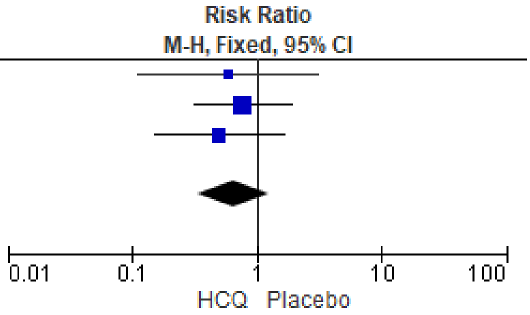


Fig. S8. Forest plot showing the risk of hospitalization for non-hospitalized COVID-19 patients treated with hydroxychloroquine.

| Study or Subgroup | HCQ Events | HCQ Total | Placebo Events | Placebo Total | Weight | Risk Ratio M-H, Fixed, 95% CI |
|---|---|---|---|---|---|---|
| Johnston 2021 | 0 | 71 | 0 | 83 | | Not estimable |
| Reis 2021 | 0 | 214 | 1 | 227 | 59.2% | 0.35 [0.01, 8.63] |
| Skipper 2020 | 1 | 212 | 1 | 211 | 40.8% | 1.00 [0.06, 15.81] |
| **Total (95% CI)** | | **497** | | **521** | **100.0%** | **0.62 [0.08, 4.68]** |
| Total events | 1 | | 2 | | | |

Heterogeneity: Chi² = 0.23, df = 1 (P = 0.63); I² = 0%
Test for overall effect: Z = 0.47 (P = 0.64)

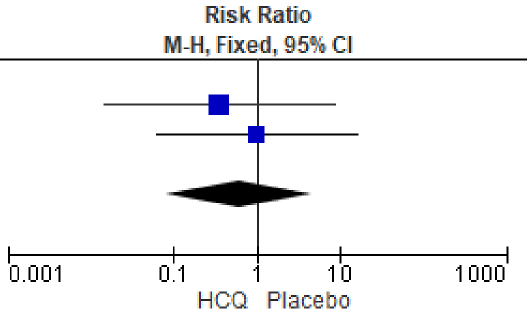


Fig. S9. Forest plot showing the risk of death for non-hospitalized COVID-19 patients treated with hydroxychloroquine.

| Study or Subgroup | HCQ Events | HCQ Total | Placebo Events | Placebo Total | Weight | Risk Ratio M-H, Random, 95% CI |
|---|---|---|---|---|---|---|
| Johnston 2021 | 5 | 71 | 5 | 83 | 13.6% | 1.17 [0.35, 3.88] |
| Reis 2021 | 46 | 207 | 46 | 220 | 41.7% | 1.06 [0.74, 1.53] |
| Skipper 2020 | 92 | 212 | 46 | 211 | 44.7% | 1.99 [1.48, 2.68] |
| **Total (95% CI)** | | **490** | | **514** | **100.0%** | **1.43 [0.85, 2.38]** |
| Total events | 143 | | 97 | | | |

Heterogeneity: Tau² = 0.13; Chi² = 7.08, df = 2 (P = 0.03); I² = 72%
Test for overall effect: Z = 1.35 (P = 0.18)

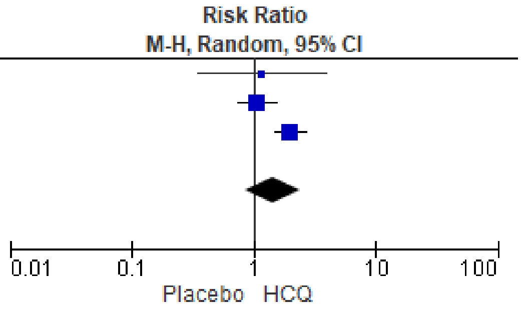


Fig. S10. Forest plot showing the risk of any adverse events for non-hospitalized COVID-19 patients treated with hydroxychloroquine.

Table S8. Blinded, placebo-controlled, randomized clinical trials evaluating the efficacy and safety of hydroxychloroquine as treatment for hospitalized patients with COVID-19.

| Author | Study | Country | Protocol and Phase | Population | Intervention | Control | Outcomes of interest (Time of evaluation) |
|---|---|---|---|---|---|---|---|
| Dubée 2021 | Hydroxychloroquine in mild-to-moderate COVID-19: a placebo-controlled double-blind trial | France | NCT04325893 (Phase 3) | Non-critically ill patients with COVID-19 requiring hospitalization with/without need for supplemental oxygen. Cases requiring oxygen therapy >3 L/min or intensive care were excluded.<br>• HCQ group: median age 76y (range 60-85). Sex: 52.0% male. Medical conditions: hypertension 52.0%, diabetes 16.0%, heart disease 27.2%. Concomitant medications: corticosteroids 10.4%.<br>• Placebo: median age 78y (range 57-87). Sex: 44.8% male. Medical conditions: hypertension 54.8%, diabetes 18.6%, heart disease 30.7%. Concomitant medications: corticosteroids 8.8%. | HCQ 200mg, 2x/day in the first day + 200mg, 2x/day for 8 days (n = 124) | Placebo (n = 123) | Need for mechanical ventilation, death, and adverse events (14 and 28 days) |
| Hernandez-Cardenas 2021 | Hydroxychloroquine for the treatment of severe respiratory infection by COVID-19: a randomized controlled trial | Mexico | NCT04315896 (Phase 3) | Critically ill patients with COVID-19 requiring hospitalization with/without need for mechanical ventilation.<br>• HCQ group: mean age 50y (SD 11). Sex: 82.1% male. Medical conditions: hypertension 16.0%, diabetes 18.9%. Concomitant medications: corticosteroids 51.9%, anticoagulants 54.7%, tocilizumab 2.8%.<br>• Placebo: mean age 49y (SD 11). Sex: 68.5% male. Medical conditions: hypertension: 17.6%, diabetes 13.0%. Concomitant medications: corticosteroids 48.1%, anticoagulants 57.4%, tocilizumab 1.9%. | HCQ 200mg, 2x/day for 10 days (n = 106) | Placebo (n = 108) | Days of hospitalization, death, and adverse events (30 days) |
| Gonzalez 2021 | Efficacy and safety of Ivermectin and Hydroxychloroquine in patients with severe COVID-19. A randomized controlled trial | Mexico | NCT04391127 (Phase 3) | Non-critically ill patients with COVID-19 requiring hospitalization with need for supplemental oxygen. Cases requiring oxygen therapy >10 L/min or mechanical ventilation were excluded.<br>• HCQ group: mean age 48.9y (SD 15.3). Sex: 66.6% male. Medical conditions: hypertension 24.2%, diabetes 33.3%. Concomitant medications: corticosteroids 63.6%, anticoagulants 90.9%.<br>• Placebo: mean age 53.8y (SD 16.9). Sex: 62.1% male. Medical conditions: hypertension 37.8%, diabetes 43.2%. Concomitant medications: corticosteroids 51.3%, anticoagulants 94.5%. | HCQ 400mg, 2x/day in the first day + 200mg, 2x/day for 4 days (n = 33) | Placebo (n = 37) | Days of hospitalization, death, and adverse events (30 days) |

Table S8 (continuation). Blinded, placebo-controlled, randomized clinical trials evaluating the efficacy and safety of hydroxychloroquine as treatment for hospitalized patients with COVID-19.

| Author | Study | Country | Protocol and Phase | Population | Intervention | Control | Outcomes of interest (Time of evaluation) |
|---|---|---|---|---|---|---|---|
| Ulrich 2020 | Treating COVID-19 With Hydroxychloroquine (TEACH): A Multicenter, Double-Blind Randomized Controlled Trial in Hospitalized Patients | USA | NCT04369742 (Phase 2) | Non-critically ill patients with COVID-19 requiring hospitalization with/without need for supplemental oxygen. Cases requiring mechanical ventilation or intensive care on enrollment were excluded.<br>• HCQ group: mean age 66.5y (SD 16.4). Sex: 67.2% male. Medical conditions: hypertension 53.7%, diabetes 28.4%, heart disease 31.3%. Concomitant medications: corticosteroids 10.4%, anticoagulants 32.8%, remdesivir 1.5%, tocilizumab 4.5%.<br>• Placebo: mean age 65.8y (SD 16.0). Sex: 50.8% male. Medical conditions: hypertension 62.3%, diabetes 36.1%. Concomitant medications: corticosteroids 9.8%, anticoagulants 39.8%, remdesivir 0, tocilizumab 3.3%. | HCQ 400mg, 2x/day in the first day + 200mg, 2x/day for 4 days (n = 67) | Placebo (n = 61) | Need for mechanical ventilation, days of hospitalization, death, and adverse events (14 and 30 days) |
| Self 2020 | Effect of Hydroxychloroquine on Clinical Status at 14 Days in Hospitalized Patients With COVID-19: A Randomized Clinical Trial | USA | NCT04332991 (Phase 3) | Critically and non-critically patients with COVID-19 requiring hospitalization with/without intensive care.<br>• HCQ group: median age 58y (range 45-69). Sex: 55.8% male. Medical conditions: hypertension 56.2%, diabetes 36.4%, heart disease 7.9%. Concomitant medications: corticosteroids 16.1%, remdesivir 23.1%, tocilizumab 4.1%.<br>• Placebo: median age 57y (range 43-68). Sex: 55.7% male. Medical conditions: hypertension 49.4%, diabetes 32.9%, heart disease 9.7%. Concomitant medications: corticosteroids 20.7%, remdesivir 20.3%, tocilizumab 7.2%. | HCQ 400mg, 2x/day in the first day + 200mg, 2x/day for 4 days (n = 242) | Placebo (n = 237) | Need for mechanical ventilation, death, and adverse events (14 and 28 days) |

HCQ, hydroxychloroquine.

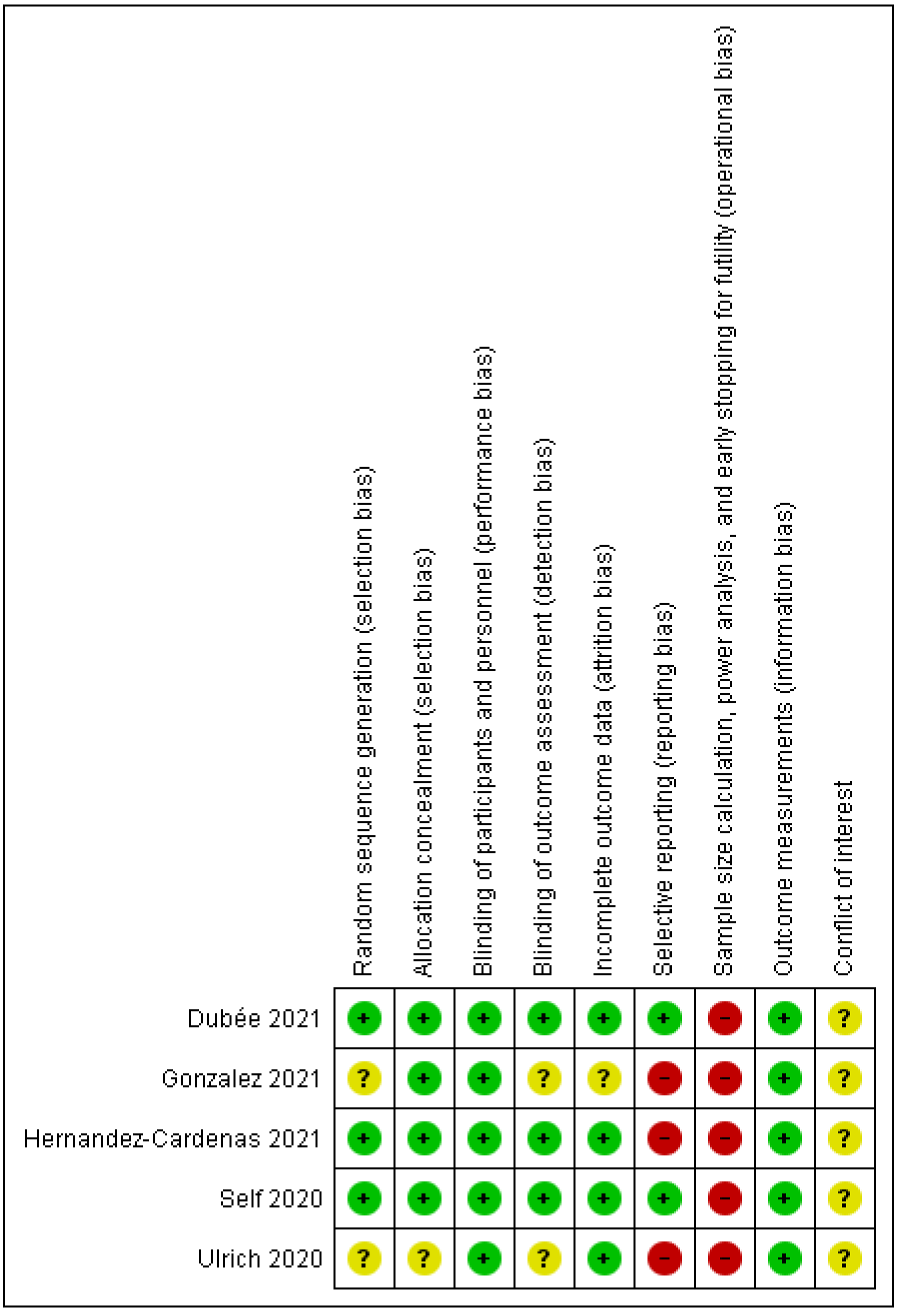

Fig. S11. Risk of bias assessment on the use of hydroxychloroquine as treatment for hospitalized patients with COVID-19.

Table S9. Individual results of studies evaluating the efficacy and safety of hydroxychloroquine as treatment for hospitalized patients with COVID-19.

| Author | Days of hospitalization | Mechanical ventilation | Death | Any adverse events | Conclusion |
|---|---|---|---|---|---|
| Dubée 2021 | - | 14 days<br>HCQ: 3/124 (2.4%)<br>Placebo: 3/123 (2.4%)<br><br>28 days<br>HCQ: 3/124 (2.4%)<br>Placebo: 4/123 (3.3%) | 14 days<br>HCQ: 6/124 (4.8%)<br>Placebo: 6/123 (4.9%)<br><br>28 days<br>HCQ: 6/124 (4.8%)<br>Placebo: 11/123 (8.9%) | 28 days<br>HCQ: 70/124 (56.5%)<br>Placebo: 61/120 (50.8%) | "Patients treated with hydroxychloroquine did not experience better clinical or virological outcomes than those receiving the placebo." |
| Hernandez-Cardenas 2021 | HCQ: 17.8 ± 11.0 days*<br>Placebo: 18.1 ± 12.0 days | - | 30 days<br>HCQ: 40/106 (37.7%)<br>Placebo: 44/108 (40.7%) | Data on adverse events were unclear and could not be extracted | "No beneficial effect or significant harm could be demonstrated in our randomized controlled trial including 214 patients, using relatively low doses of HCQ compared with placebo in hospitalized patients with severe COVID-19." |
| Gonzalez 2021 | HCQ: 7 (3 – 9) days**<br>Placebo: 5 (4 –7) days | - | 30 days<br>HCQ: 2/33 (6.1%)<br>Placebo: 6/37 (16.2%) | Data on adverse events were unclear and could not be extracted | "In non-critical hospitalized patients with COVID-19 pneumonia, neither ivermectin nor hydroxychloroquine decreases the number of in-hospital days, respiratory deterioration, or deaths." |
| Ulrich 2020 | HCQ: 9.8 ± 10.3 days*<br>Placebo: 6.8 ± 5.9 days | 14 days<br>HCQ: 5/67 (7.5%)<br>Placebo: 4/61 (6.6%)<br><br>30 days<br>HCQ: 5/67 (7.5%)<br>Placebo: 3/61 (4.9%) | 14 days<br>HCQ: 3/67 (4.5%)<br>Placebo: 5/61 (8.2%)<br><br>30 days<br>HCQ: 7/67 (10.4%)<br>Placebo: 6/61 (9.8%) | 30 days<br>HCQ: 38/67 (56.7%)<br>Placebo: 36/61 (59.0%) | "In hospitalized patients with COVID-19, our data suggest that HCQ does not prevent severe outcomes or improve clinical scores." |
| Self 2020 | - | 14 days<br>HCQ: 18/242 (7.4%)<br>Placebo: 24/237 (10.1%)<br><br>28 days<br>HCQ: 11/242 (4.5%)<br>Placebo: 12/237 (5.1%) | 14 days<br>HCQ: 18/242 (7.4%)<br>Placebo: 14/237 (5.9%)<br><br>28 days<br>HCQ: 25/242 (10.3%)<br>Placebo: 25/237 (10.5%) | 28 days<br>HCQ: 14/242 (5.8%)<br>Placebo: 11/237 (4.6%) | "These findings do not support the use of hydroxychloroquine for treatment of COVID-19 among hospitalized adults." |

HCQ, hydroxychloroquine. * Data were reported in mean and standard deviation. ** Data were reported in median and interquartile range.

| Study or Subgroup | HCQ Events | HCQ Total | Placebo Events | Placebo Total | Weight | Risk Ratio M-H, Fixed, 95% CI |
|---|---|---|---|---|---|---|
| Dubée 2021 | 3 | 124 | 3 | 123 | 9.6% | 0.99 [0.20, 4.82] |
| Self 2020 | 18 | 242 | 24 | 237 | 77.1% | 0.73 [0.41, 1.32] |
| Ulrich 2020 | 5 | 67 | 4 | 61 | 13.3% | 1.14 [0.32, 4.05] |
| **Total (95% CI)** | | **433** | | **421** | **100.0%** | **0.81 [0.49, 1.34]** |
| Total events | 26 | | 31 | | | |

Heterogeneity: Chi$^2$ = 0.45, df = 2 (P = 0.80); $I^2$ = 0%
Test for overall effect: Z = 0.81 (P = 0.42)

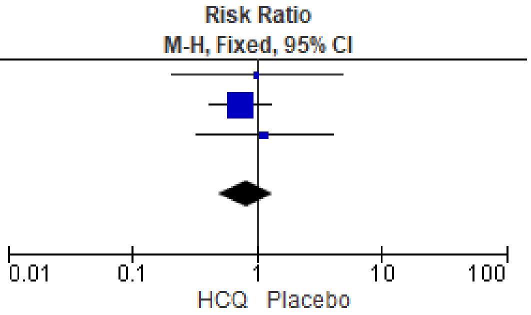


Fig. S12. Forest plot showing the risk of mechanical ventilation at two weeks for hospitalized COVID-19 patients treated with hydroxychloroquine.

| Study or Subgroup | HCQ Events | HCQ Total | Placebo Events | Placebo Total | Weight | Risk Ratio M-H, Fixed, 95% CI |
|---|---|---|---|---|---|---|
| Dubée 2021 | 3 | 124 | 4 | 123 | 20.8% | 0.74 [0.17, 3.26] |
| Self 2020 | 11 | 242 | 12 | 237 | 62.9% | 0.90 [0.40, 1.99] |
| Ulrich 2020 | 5 | 67 | 3 | 61 | 16.3% | 1.52 [0.38, 6.08] |
| **Total (95% CI)** | | **433** | | **421** | **100.0%** | **0.97 [0.52, 1.80]** |
| Total events | 19 | | 19 | | | |

Heterogeneity: Chi$^2$ = 0.56, df = 2 (P = 0.76); $I^2$ = 0%
Test for overall effect: Z = 0.11 (P = 0.91)

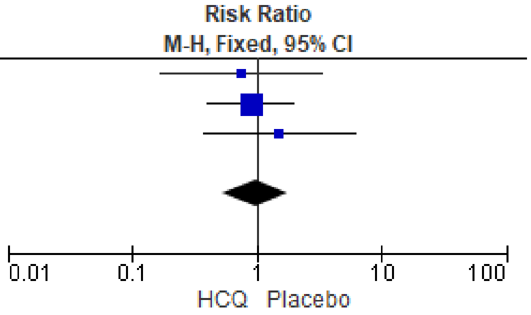


Fig. S13. Forest plot showing the risk of mechanical ventilation at four weeks for hospitalized COVID-19 patients treated with hydroxychloroquine.

| Study or Subgroup | HCQ Events | HCQ Total | Placebo Events | Placebo Total | Weight | Risk Ratio M-H, Fixed, 95% CI |
|---|---|---|---|---|---|---|
| Dubée 2021 | 6 | 124 | 6 | 123 | 23.7% | 0.99 [0.33, 2.99] |
| Self 2020 | 18 | 242 | 14 | 237 | 55.7% | 1.26 [0.64, 2.47] |
| Ulrich 2020 | 3 | 67 | 5 | 61 | 20.6% | 0.55 [0.14, 2.19] |
| **Total (95% CI)** | | **433** | | **421** | **100.0%** | **1.05 [0.62, 1.78]** |
| Total events | 27 | | 25 | | | |

Heterogeneity: Chi$^2$ = 1.14, df = 2 (P = 0.57); $I^2$ = 0%
Test for overall effect: Z = 0.18 (P = 0.86)

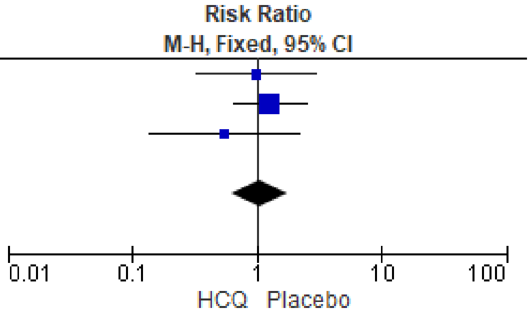


Fig. S14. Forest plot showing the risk of death at two weeks for hospitalized COVID-19 patients treated with hydroxychloroquine.

| Study or Subgroup | HCQ Events | HCQ Total | Placebo Events | Placebo Total | Weight | Risk Ratio M-H, Fixed, 95% CI |
|---|---|---|---|---|---|---|
| Dubée 2021 | 6 | 124 | 11 | 123 | 12.0% | 0.54 [0.21, 1.42] |
| Gonzalez 2021 | 2 | 33 | 6 | 37 | 6.2% | 0.37 [0.08, 1.73] |
| Hernandez-Cardenas 2021 | 40 | 106 | 44 | 108 | 47.5% | 0.93 [0.66, 1.29] |
| Self 2020 | 25 | 242 | 25 | 237 | 27.5% | 0.98 [0.58, 1.65] |
| Ulrich 2020 | 7 | 67 | 6 | 61 | 6.8% | 1.06 [0.38, 2.99] |
| **Total (95% CI)** | | **572** | | **566** | **100.0%** | **0.87 [0.67, 1.13]** |
| Total events | 80 | | 92 | | | |

Heterogeneity: Chi$^2$ = 2.58, df = 4 (P = 0.63); $I^2$ = 0%
Test for overall effect: Z = 1.04 (P = 0.30)

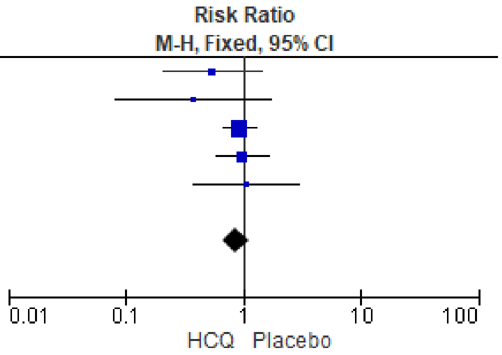


Fig. S15. Forest plot showing the risk of death at four weeks for hospitalized COVID-19 patients treated with hydroxychloroquine.

| Study or Subgroup | HCQ Events | HCQ Total | Placebo Events | Placebo Total | Weight | Risk Ratio M-H, Fixed, 95% CI |
|---|---|---|---|---|---|---|
| Dubée 2021 | 70 | 124 | 61 | 120 | 56.0% | 1.11 [0.88, 1.40] |
| Self 2020 | 14 | 242 | 11 | 237 | 10.0% | 1.25 [0.58, 2.69] |
| Ulrich 2020 | 38 | 67 | 36 | 61 | 34.0% | 0.96 [0.71, 1.29] |
| **Total (95% CI)** | | **433** | | **418** | **100.0%** | **1.07 [0.89, 1.29]** |
| Total events | 122 | | 108 | | | |

Heterogeneity: Chi² = 0.76, df = 2 (P = 0.68); I² = 0%
Test for overall effect: Z = 0.75 (P = 0.45)

Risk Ratio
M-H, Fixed, 95% CI

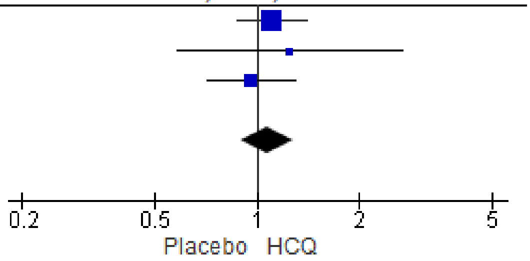


Fig. S16. Forest plot showing the risk of any adverse events for hospitalized COVID-19 patients treated with hydroxychloroquine.

Table S10. Sensitivity analysis of studies evaluating the efficacy and safety of hydroxychloroquine as treatment for hospitalized patients with COVID-19.

| Outcomes | RR (95% CI) | $I^2$ | Risk of bias | Inconsistency | Indirectness | Imprecision | Influence of small trials | Large effect | Quality of evidence |
|---|---|---|---|---|---|---|---|---|---|
| Death (4 weeks) | | | | | | | | | |
| Peer-reviewed studies | 0.88 (0.58 to 1.33) | 0% | Not serious | Not serious | Not serious | Serious | Not serious | No | ⊕⊕⊕◯ |
| Non-peer reviewed studies | 0.80 (0.42 to 1.55) | 24% | Not serious | Not serious | Not serious | Serious | Not serious | No | ⊕⊕⊕◯ |

Certainty: ⊕ very-low; ⊕⊕ low; ⊕⊕⊕ moderate; ⊕⊕⊕⊕ high.

### Risk of adverse events for individuals using hydroxychloroquine compared to placebo

| Study or Subgroup | HCQ Events | HCQ Total | Placebo Events | Placebo Total | Weight | Risk Ratio M-H, Random, 95% CI |
|---|---|---|---|---|---|---|
| Abella 2021 | 29 | 65 | 17 | 65 | 7.7% | 1.71 [1.04, 2.78] |
| Barnabas 2020 | 66 | 407 | 46 | 422 | 9.6% | 1.49 [1.05, 2.11] |
| Boulware 2020 | 140 | 349 | 59 | 351 | 10.8% | 2.39 [1.83, 3.11] |
| Dubée 2021 | 70 | 124 | 61 | 120 | 11.3% | 1.11 [0.88, 1.40] |
| Johnston 2021 | 5 | 71 | 5 | 83 | 2.6% | 1.17 [0.35, 3.88] |
| Rajasingham 2020 | 316 | 936 | 100 | 469 | 11.7% | 1.58 [1.30, 1.93] |
| Reis 2021 | 46 | 207 | 46 | 220 | 9.5% | 1.06 [0.74, 1.53] |
| Rojas-Serrano 2021 | 32 | 62 | 38 | 65 | 10.1% | 0.88 [0.64, 1.21] |
| Self 2020 | 14 | 242 | 11 | 237 | 4.9% | 1.25 [0.58, 2.69] |
| Skipper 2020 | 92 | 212 | 46 | 211 | 10.4% | 1.99 [1.48, 2.68] |
| Syed 2021 | 9 | 154 | 1 | 46 | 1.0% | 2.69 [0.35, 20.67] |
| Ulrich 2020 | 38 | 67 | 36 | 61 | 10.4% | 0.96 [0.71, 1.29] |
| **Total (95% CI)** | | **2896** | | **2350** | **100.0%** | **1.38 [1.12, 1.71]** |
| Total events | 857 | | 466 | | | |

Heterogeneity: Tau² = 0.09; Chi² = 45.71, df = 11 (P < 0.00001); I² = 76%
Test for overall effect: Z = 2.97 (P = 0.003)

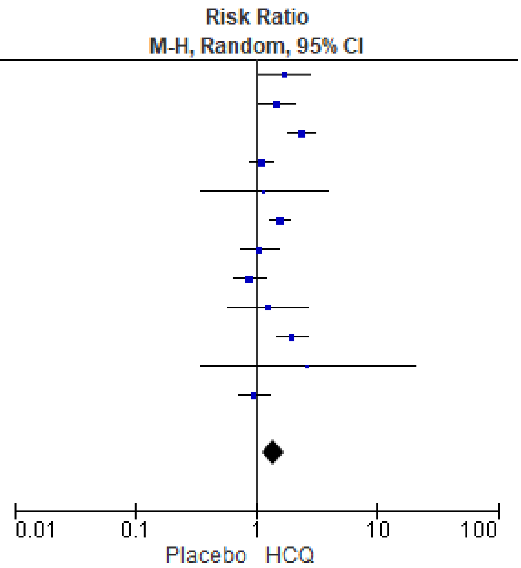


Fig. S17. Forest plot showing the risk of any adverse events for patients treated with hydroxychloroquine.

| Study or Subgroup | HCQ Events | HCQ Total | Placebo Events | Placebo Total | Weight | Risk Ratio M-H, Fixed, 95% CI |
|---|---|---|---|---|---|---|
| Barnabas 2020 | 1 | 407 | 0 | 422 | 1.4% | 3.11 [0.13, 76.13] |
| Dubée 2021 | 3 | 124 | 4 | 120 | 11.4% | 0.73 [0.17, 3.17] |
| Reis 2021 | 11 | 207 | 12 | 220 | 32.6% | 0.97 [0.44, 2.16] |
| Self 2020 | 14 | 242 | 11 | 237 | 31.2% | 1.25 [0.58, 2.69] |
| Ulrich 2020 | 9 | 67 | 8 | 61 | 23.5% | 1.02 [0.42, 2.49] |
| **Total (95% CI)** | | **1047** | | **1060** | **100.0%** | **1.07 [0.69, 1.67]** |
| Total events | 38 | | 35 | | | |

Heterogeneity: Chi² = 0.91, df = 4 (P = 0.92); I² = 0%
Test for overall effect: Z = 0.31 (P = 0.76)

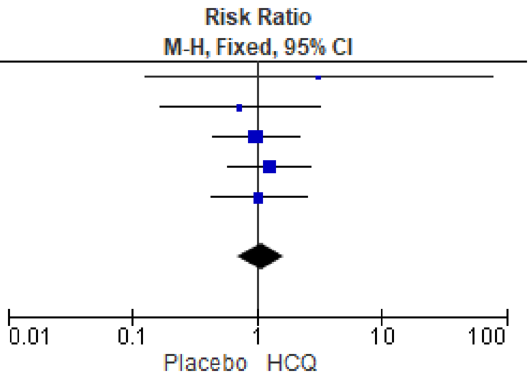


Fig. S18. Forest plot showing the risk of serious adverse events for patients treated with hydroxychloroquine.

| Study or Subgroup | HCQ Events | HCQ Total | Placebo Events | Placebo Total | Weight | Risk Ratio M-H, Fixed, 95% CI |
|---|---|---|---|---|---|---|
| Johnston 2021 | 2 | 71 | 0 | 83 | 5.4% | 5.83 [0.28, 119.53] |
| Self 2020 | 13 | 221 | 7 | 214 | 82.5% | 1.80 [0.73, 4.42] |
| Ulrich 2020 | 3 | 67 | 1 | 61 | 12.1% | 2.73 [0.29, 25.57] |
| **Total (95% CI)** | | **359** | | **358** | **100.0%** | **2.13 [0.96, 4.71]** |
| Total events | 18 | | 8 | | | |

Heterogeneity: Chi² = 0.61, df = 2 (P = 0.74); I² = 0%
Test for overall effect: Z = 1.86 (P = 0.06)

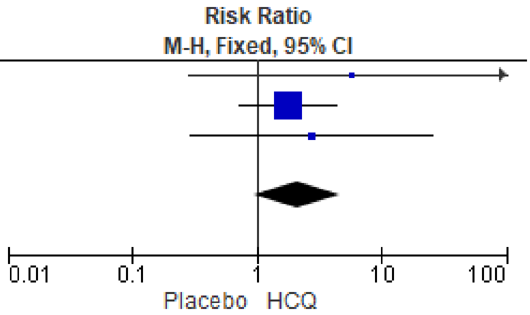


Fig. S19. Forest plot showing the risk of QTc interval > 500ms for patients treated with hydroxychloroquine.

| Study or Subgroup | HCQ Events | HCQ Total | Placebo Events | Placebo Total | Weight | Risk Ratio M-H, Random, 95% CI |
|---|---|---|---|---|---|---|
| Dubée 2021 | 6 | 124 | 7 | 120 | 24.1% | 0.83 [0.29, 2.40] |
| Johnston 2021 | 19 | 71 | 2 | 83 | 19.2% | 11.11 [2.68, 46.04] |
| Reis 2021 | 0 | 207 | 2 | 220 | 7.4% | 0.21 [0.01, 4.40] |
| Rojas-Serrano 2021 | 3 | 62 | 3 | 65 | 17.6% | 1.05 [0.22, 5.00] |
| Self 2020 | 30 | 242 | 21 | 237 | 31.7% | 1.40 [0.83, 2.37] |
| **Total (95% CI)** | | **706** | | **725** | **100.0%** | **1.52 [0.60, 3.83]** |
| Total events | 58 | | 35 | | | |

Heterogeneity: Tau² = 0.63; Chi² = 11.07, df = 4 (P = 0.03); I² = 64%
Test for overall effect: Z = 0.89 (P = 0.37)

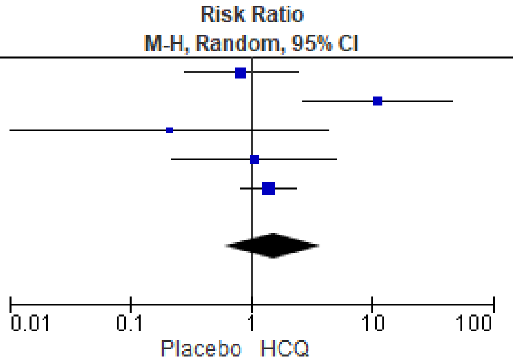


Fig. S20. Forest plot showing the risk of cardiac manifestations for patients treated with hydroxychloroquine.

| Study or Subgroup | HCQ Events | HCQ Total | Placebo Events | Placebo Total | Weight | Risk Ratio M-H, Random, 95% CI |
|---|---|---|---|---|---|---|
| Abella 2021 | 25 | 65 | 8 | 65 | 10.8% | 3.13 [1.52, 6.41] |
| Barnabas 2020 | 25 | 407 | 7 | 422 | 9.2% | 3.70 [1.62, 8.47] |
| Boulware 2020 | 81 | 349 | 15 | 351 | 14.2% | 5.43 [3.19, 9.23] |
| Dubée 2021 | 14 | 124 | 11 | 120 | 10.3% | 1.23 [0.58, 2.60] |
| Rajasingham 2020 | 140 | 936 | 35 | 469 | 17.9% | 2.00 [1.41, 2.85] |
| Rojas-Serrano 2021 | 19 | 62 | 10 | 65 | 11.4% | 1.99 [1.01, 3.94] |
| Skipper 2020 | 50 | 212 | 20 | 211 | 15.2% | 2.49 [1.54, 4.03] |
| Ulrich 2020 | 17 | 67 | 10 | 61 | 11.1% | 1.55 [0.77, 3.12] |
| **Total (95% CI)** | | **2222** | | **1764** | **100.0%** | **2.45 [1.77, 3.39]** |
| Total events | 371 | | 116 | | | |

Heterogeneity: Tau² = 0.12; Chi² = 16.72, df = 7 (P = 0.02); I² = 58%
Test for overall effect: Z = 5.38 (P < 0.00001)

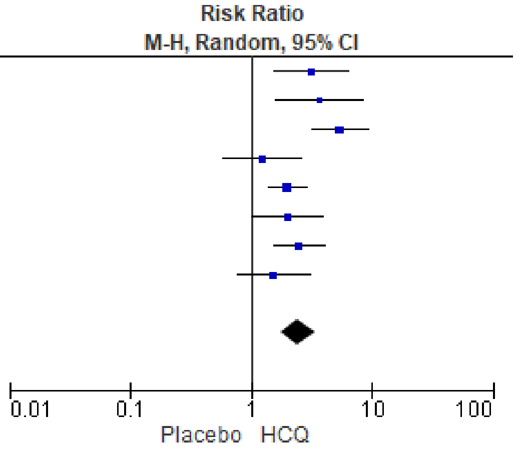


Fig. S21. Forest plot showing the risk of gastrointestinal symptoms for patients treated with hydroxychloroquine.

| Study or Subgroup | HCQ Events | HCQ Total | Placebo Events | Placebo Total | Weight | Risk Ratio M-H, Random, 95% CI |
|---|---|---|---|---|---|---|
| Abella 2021 | 0 | 65 | 4 | 65 | 3.1% | 0.11 [0.01, 2.02] |
| Barnabas 2020 | 5 | 407 | 7 | 422 | 18.4% | 0.74 [0.24, 2.31] |
| Boulware 2020 | 13 | 349 | 8 | 351 | 29.4% | 1.63 [0.69, 3.89] |
| Dubée 2021 | 3 | 124 | 1 | 120 | 5.2% | 2.90 [0.31, 27.52] |
| Johnston 2021 | 1 | 71 | 0 | 83 | 2.6% | 3.50 [0.14, 84.59] |
| Rojas-Serrano 2021 | 8 | 62 | 14 | 65 | 33.8% | 0.60 [0.27, 1.33] |
| Skipper 2020 | 2 | 212 | 0 | 121 | 2.9% | 2.86 [0.14, 59.17] |
| Ulrich 2020 | 1 | 67 | 2 | 61 | 4.6% | 0.46 [0.04, 4.90] |
| **Total (95% CI)** | | **1357** | | **1288** | **100.0%** | **0.93 [0.55, 1.56]** |
| Total events | 33 | | 36 | | | |

Heterogeneity: Tau² = 0.04; Chi² = 7.53, df = 7 (P = 0.38); I² = 7%
Test for overall effect: Z = 0.27 (P = 0.79)

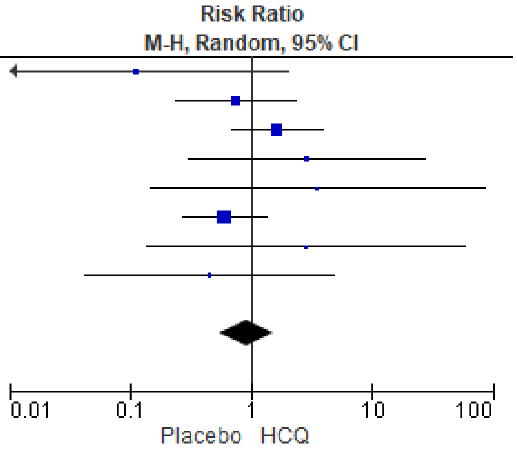


Fig. S22. Forest plot showing the risk of headache for patients treated with hydroxychloroquine.

| Study or Subgroup | HCQ Events | HCQ Total | Placebo Events | Placebo Total | Weight | Risk Ratio M-H, Fixed, 95% CI |
|---|---|---|---|---|---|---|
| Abella 2021 | 3 | 65 | 1 | 65 | 3.9% | 3.00 [0.32, 28.09] |
| Barnabas 2020 | 11 | 407 | 8 | 422 | 30.6% | 1.43 [0.58, 3.51] |
| Boulware 2020 | 4 | 349 | 2 | 351 | 7.8% | 2.01 [0.37, 10.91] |
| Dubée 2021 | 10 | 124 | 7 | 120 | 27.7% | 1.38 [0.54, 3.51] |
| Rojas-Serrano 2021 | 1 | 62 | 0 | 65 | 1.9% | 3.14 [0.13, 75.72] |
| Self 2020 | 1 | 242 | 1 | 237 | 3.9% | 0.98 [0.06, 15.57] |
| Skipper 2020 | 6 | 212 | 2 | 211 | 7.8% | 2.99 [0.61, 14.63] |
| Ulrich 2020 | 1 | 67 | 4 | 61 | 16.3% | 0.23 [0.03, 1.98] |
| **Total (95% CI)** | | **1528** | | **1532** | **100.0%** | **1.46 [0.89, 2.39]** |
| Total events | 37 | | 25 | | | |

Heterogeneity: Chi² = 4.47, df = 7 (P = 0.72); I² = 0%
Test for overall effect: Z = 1.51 (P = 0.13)

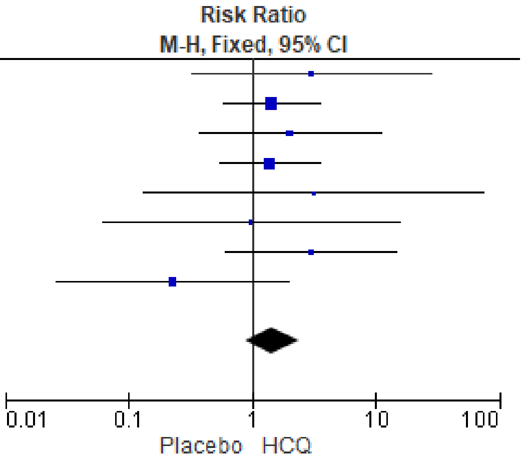


Fig. S23. Forest plot showing the risk of skin rash for patients treated with hydroxychloroquine.

| Study or Subgroup | HCQ Events | HCQ Total | Placebo Events | Placebo Total | Weight | Risk Ratio M-H, Fixed, 95% CI |
|---|---|---|---|---|---|---|
| Abella 2021 | 1 | 65 | 0 | 65 | 1.2% | 3.00 [0.12, 72.31] |
| Barnabas 2020 | 6 | 407 | 11 | 422 | 25.5% | 0.57 [0.21, 1.51] |
| Boulware 2020 | 19 | 349 | 13 | 351 | 30.7% | 1.47 [0.74, 2.93] |
| Dubée 2021 | 0 | 124 | 2 | 120 | 6.0% | 0.19 [0.01, 3.99] |
| Rojas-Serrano 2021 | 2 | 62 | 2 | 65 | 4.6% | 1.05 [0.15, 7.21] |
| Self 2020 | 1 | 242 | 0 | 237 | 1.2% | 2.94 [0.12, 71.77] |
| Skipper 2020 | 20 | 212 | 13 | 211 | 30.8% | 1.53 [0.78, 3.00] |
| **Total (95% CI)** | | **1461** | | **1471** | **100.0%** | **1.20 [0.80, 1.78]** |
| Total events | 49 | | 41 | | | |

Heterogeneity: Chi² = 5.11, df = 6 (P = 0.53); I² = 0%
Test for overall effect: Z = 0.89 (P = 0.38)

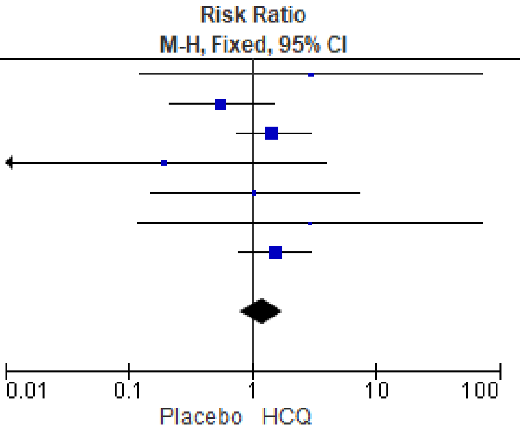


Fig. S24. Forest plot showing the risk of neurologic reactions for patients treated with hydroxychloroquine.
